# Quantitative Microscopy in Medicine

**DOI:** 10.1101/2024.07.31.24311304

**Authors:** Alexandre Matov

## Abstract

**Introduction:** Methods for personalizing medical treatment are the focal point of contemporary biomedical research. In cancer care, we can analyze the effects of therapies at the level of individual cells. Quantitative characterization of treatment efficacy and evaluation of why some individuals respond to specific regimens, whereas others do not, requires additional approaches to genetic sequencing at single time points. Methods for the analysis of changes in phenotype, such as *in vivo* and *ex vivo* morphology and localization of cellular proteins and organelles can provide important insights into patient treatment options.

**Methods:** Novel therapies are needed to extend survival in metastatic castration-resistant prostate cancer (mCRPC). Prostate-specific membrane antigen (PSMA), a cell surface glycoprotein that is commonly overexpressed by prostate cancer (PC) cells relative to normal prostate cells, provides a validated target.

**Results:** We developed a software for image analysis designed to identify PSMA expression on the surface of epithelial cells in order to extract prognostic metrics. In addition, our software can deliver predictive information and inform clinicians regarding the efficacy of PC therapy. We can envisage additional applications of our software system, beyond PC, as PSMA is expressed in a variety of tissues. Our method is based on image denoising, topologic partitioning, and edge detection. These three steps allow to segment the area of each PSMA spot in an image of a coverslip with epithelial cells.

**Conclusions:** Our objective has been to present the community with an integrated, easy to use by all, tool for resolving the complex cellular organization and it is our goal to have such software system approved for use in the clinical practice.

## INTRODUCTION

Prostate cancer (PC) affects hundreds of thousands of men each year in the United States and around the World. Despite advances in diagnostic and treatment strategies, PC is the second most common cause of cancer mortality in men (Bray et al., 2018). Though treatment options for metastatic castration-resistant PC (mCRPC) have expanded over the past decade, highly proliferative phenotypes frequently emerge at the time of progression on androgen signaling inhibitors (Tannock et al., 2004). Though initially effective in reducing tumor burden for some patients, resistance to systemic is universal and approximately one-third of tumors are primarily refractory to this treatment approach (Petrylak et al., 2004). This is clinically highly significant as mCRPC refractory to tubulin inhibitors is uniformly fatal within 12-18 months (Francini et al., 2019). Currently, mCRPC chemotherapy is limited to FDA-approved docetaxel and cabazitaxel, and could be potentially impactful (Sung et al., 2013; Sung et al., 2012). However, even for the FDA-approved taxanes, there is currently no good mechanistic understanding of drug action and, for instance, docetaxel is always used as a first-line chemotherapy without having to examine patient cells *ex vivo* for susceptibility. For patients with progressive mCRPC that was refractory to androgen receptor (AR) pathway inhibitors and had received or been deemed ineligible for microtubule (MT)-targeting taxane chemotherapy, there are not many treatment options available (Thadani-Mulero et al., 2014). For patients with mutated BRCA1/2, treatment with olaparib (and other PARP inhibitors) offers a benefit (De Santis et al., 2024; Mateo et al., 2024) and novel therapies are needed to extend survival (Unterrainer et al., 2024) in patients with visceral (especially hepatic) metastases.

Prostate-specific membrane antigen (PSMA), a cell surface antigen overexpressed in PC, provides a validated target (Bander et al., 2005). ^225^Ac-J591, anti-PSMA monoclonal antibody J591 radiolabeled with the alpha emitter actinium-225 is being investigated in the context of safety, efficacy, maximum tolerated dose (MTD), and recommended for phase II dose (RP2D) (Tagawa et al., 2024). PSMA expression is found in higher-grade PC as well as mCRPC (Bostwick et al., 1998), while its expression is low in normal tissues (Silver et al., 1997), and recent clinical trials have validated PSMA as a target for therapy for high-risk and advanced PC (Sartor et al., 2021), demonstrating it may offer benefits over the use of tubulin inhibitors (Hofman et al., 2021). This highlights the need to develop methods to inform PSMA-targeting therapeutic strategies, which is the focus of this contribution.

Late stage solid tumors of the prostate (and the breast) are associated with a high degree of morbidity and mortality. Treatment with tubulin inhibitors is a last resort, but the success is hampered by the lack of diagnostic and prognostic markers to aid physicians monitor treatment efficacy. Assays to detect circulating tumor cells (CTCs) in the peripheral blood of cancer patients before and during treatment have been used clinically to provide prognostic information, but the robustness of their CTC isolation and enumeration is limited. Optimizing CTC detection, via the utilization of our computer vision algorithms, will result in a more accurate CTC count, as a first indication of disease progression and treatment response. Importantly, the technology we present here will also measure specific CTC parameters that allow us to directly monitor treatment response, that is, treatment assessing CTC parameters.

The analysis of immunofluorescence images can be accomplished with algorithms for automated feature segmentation and the computation of morphology metrics. Different computational strategies consider per-field (or per-image) and per-cell analysis approaches based on the particularities of the datasets. Approaches for extracting metrics over the full field of view can identify specific shapes, such as circles (Matov, 2024d) and lines (Mattavelli et al., 2001). Radon transform (Radon, 1917) is the basis for the beamlet (or curvelet) transform (Donoho and Duncan, 2000), which allows the detection of curvilinear structures. In noisy images, curvilinear cellular filaments and edges can be identified via a feature-adapted beamlet transform (Berlemont and Olivo-Marin, 2010). Curvilinear features can also be detected by an approach (Koh et al., 2012) based on the scale-invariant feature transform (Lowe, 1999) and entropy (Shannon, 1948) analysis. Per-field analysis can also deliver numerical metrics (Murphy et al., 2003) regarding protein texture and morphology (Haralick et al., 1973). To improve on the performance of texton-based texture classifiers (Varma and Zisserman, 2003) and Gabor histogram features (Vitaladevuni et al., 2008), radon-like features (Kumar et al., 2010) have been developed for the segmentation of cellular organelles, such as mitochondria.

Further, PC is driven by the AR activity and breast cancer is analogously driven by estrogen receptor activity. Nuclear accumulation of these steroid hormone receptors is a key indication for their activity and such analysis requires imaging the nucleus as well. We evaluated the efficacy of taxane therapy based on the ability of preventing AR nuclear localization in CTCs (Matov, 2024b) in cohorts of metastatic PC patients before and after treatment. To perform per-cell analysis, we segmented the areas belonging to the cytoplasm and the nucleus of individual cells. In the case of nuclear staining, we also tested existing methodology for finding the boundaries between adjacent DAPI regions in an image via a graph-cuts-based binarization (Boykov and Jolly, 2001) based on which cellular seed points are detected by combining multiscale Laplacian-of-Gaussian filtering (Maxwell, 1873) constrained by distance-map-based adaptive scales selection (Lindeberg, 1998). These points were used to perform an initial segmentation that was refined utilizing a second graph-cuts-based algorithm incorporating the method of alpha expansions and graph coloring to reduce computational complexity (Al-Kofahi and al., 2010). The overall accuracy of the segmentation algorithm exceeded 86% according to manual validation for 25 representative images (15 *in vitro* images and 10 *in vivo* images, containing more than 7,400 cells) drawn from diverse cancer histopathology studies. Such approaches are applicable to the segmentation of DAPI and CD45 images, but not to PSMA and cytokeratin (CK) images, because of the reduced contrast and signal-to-noise ratio in epithelial markers datasets. An additional challenge was the ability to distinguish multi-nuclei cells from overlapping nuclei of adjacent cells.

## RESULTS

### Per-field drug-target engagement metrics

We performed image analysis of the morphology and localization of the MT network in seven diffuse gastric cancer (GC) cell lines (Tan et al., 2011). Our analysis was based on the measurements of 21 image metrics related to changes in the distribution and organization of pixel intensities of the MT cytoskeleton in baseline images and after treatment with 100 nM docetaxel. The features (Boland and Murphy, 2001) describe three categories of characteristics of the MT organization – (i) size and distribution of the MTs, (ii) MT localization as it relates to the cell edge, and (iii) shape of the MT network.

The first category comprises eight features, including the (1) number of bright objects in a cell with MT labeling, (2) Euler number (Maurolico, 1537) (distinguishes reticular or mesh-like patterns), (3) number of above-threshold pixels per object (that is, object size) and (4) their variance, (5) ratio of the size of the largest to smallest object, (6) average object distance to cell center of fluorescence and (7) its variance as well as the (8) ratio of largest to smallest distance.

The second category comprises five features, including the (9) fraction of above an intensity threshold pixels along the cell edge (distinguishes MTs that localize along the cell edge), (10) homogeneity of the gradient of cell edge intensity, (11) homogeneity in the direction of the cell edge (identifies patterns containing edges oriented predominantly along a particular direction), (12) ratio of the largest to the next largest value (or bin) in the histogram of the cell edge direction, and (13) cell edge direction difference (feature distinguishing MT patterns in which there are parallel edges).

The third category comprises eight features, including the (14) average length of the morphological skeleton (Lantuéjoul, 1977) of the MT network, (15) ratio of object skeleton length to the area of the convex hull (Graham, 1972) of the skeleton, (16) fraction of object pixels contained within the skeleton, (17) fraction of object (or pixels) fluorescence contained within the skeleton, (18) ratio of the number of branch points in the morphological skeleton to the overall length of the MT network, (19) fraction of the MT network convex hull pixels above an intensity threshold, (20) roundness of the convex hull, and (21) its elongations or eccentricity.

The 21 features exhibited a different degree of variability in the seven cell lines and their distributions at baseline as well as the changes after docetaxel treatment are shown in Fig. S1, Fig. S2, and Fig. S3. The statistical analysis with significance values is presented in Table S1. According to our analysis of the changes in the MT cytoskeleton, the docetaxel-sensitive cells were SNU1, TMK1, and AZ521 – for which changes in 4 to 10 metrics were statistically significant - and the docetaxel-resistant cells were MKN7, SCH, HS746T, and MKN45 for which changes in 1 to 3 metrics were statistically significant. Our new analysis results regarding drug resistance were confirmed by IC50 value measurements in these diffuse GC cell lines (Matov, 2024i). Since diffuse GC tumors are sensitive to treatment with cisplatin, our hypothesis is that tumors with molecular characteristics similar to cell lines SNU1, TMK1, and AZ521 will recede after treatment with taxane-based chemotherapy.

### Per-cell analysis and PSMA image segmentation

To perform analysis of individual cells, we have developed an image analysis method for segmenting the cellular areas with cell surface expression of PSMA or CK. PSMA imaging has been suggested as highly sensitive modality for detection of metastases in patients with biochemically recurrent or advanced PC (Vlachostergios et al., 2021). Phenotype analysis can confirm the presence of significant cell surface expression of PSMA before initiating treatment (Tasaki et al., 2014). Further, during treatment, a determination of an effective drug-target engagement of PSMA, evidenced by changes in image metrics (Galletti et al., 2013; Galletti et al., 2014a; Matov, 2024b; Matov, 2024h), may allow early detection of molecular response, or lack thereof, to treatment. This way, our computational approach will improve the precision of therapy and its customization to the individual.

Besides analysis of PSMA, in image datasets acquired with low (10x) magnification, our analysis relies on nuclear DAPI staining to confirm the detection of a cell. In addition, we analyze CD45 leukocyte labeling to exclude white blood cells from the analysis. Additional two channels, with MT and AR labeling, are analyzed for protein morphology and localization as drug resistance biomarkers (Matov, 2024i). In many coverslips, our computational algorithm detected small CTCs with diameter 7-9 µm (Fig. 1) (Matov, 2024d). The full area of 8 mm of the coverslips was analyzed via the processing of 169 (13 x 13) tile images per channel, that is, a full dataset consists of 507 PSMA, DAPI, and CD45 images (Fig. 2, Fig. S4). The detected CTCs were validated by manual selection, with further information on algorithm benchmarking presented in (Matov, 2024f).

**Figure 1.**
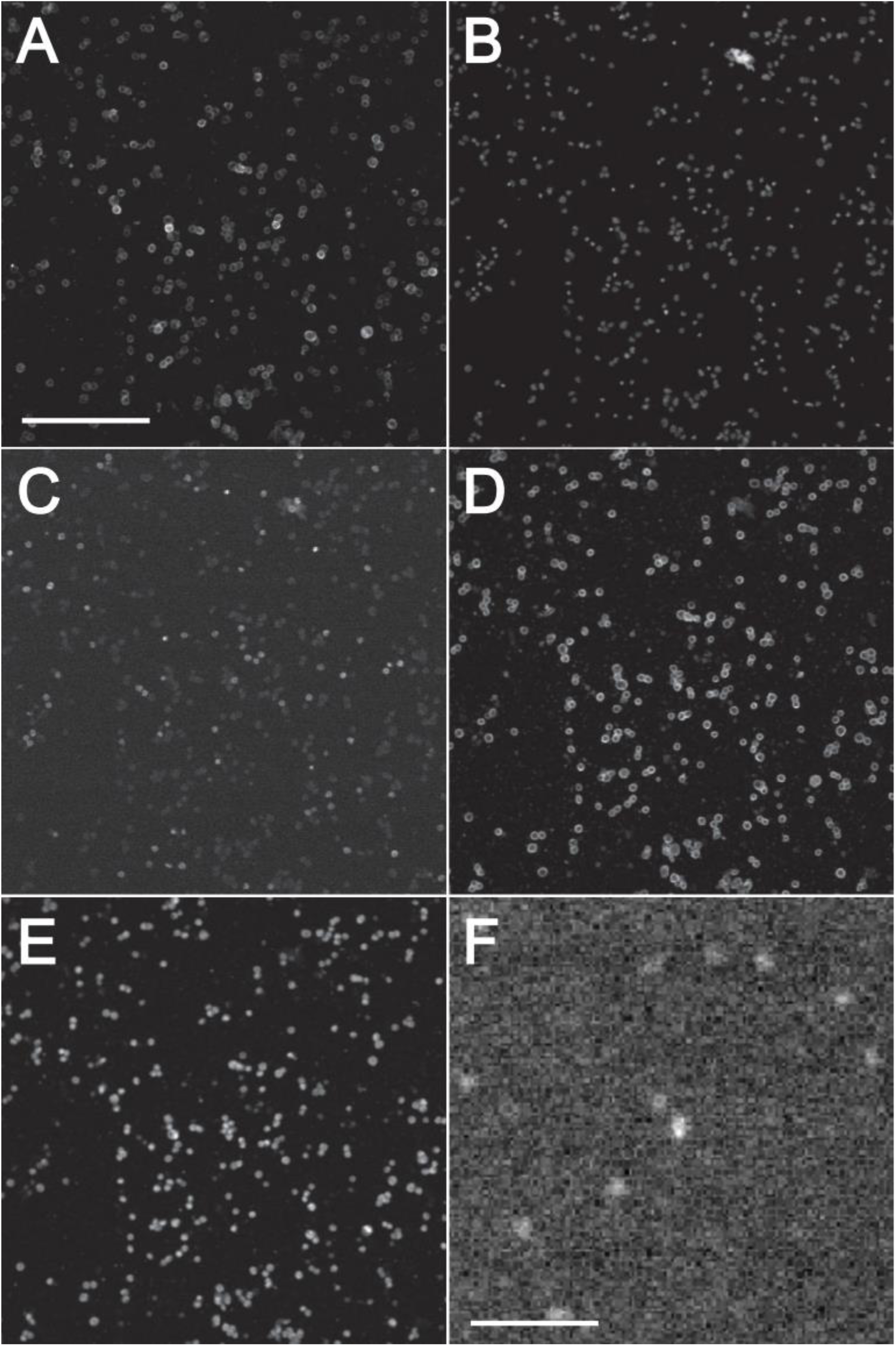
Multiplex microscopy images of patient blood. Five different markers were stained and images. Magnification, 10x. (A) Raw image of PSMA labeling. Scale bar equals 300 µm. (B) DAPI nuclear labeling. (C) CD45 leukocytes labeling. (D) MT marker. (E) AR labeling. (F) Zoom-in of computationally identified PSMA spots; the diameter of the bright spot in the middle of the image is 8.8 µm. Scale bar equals 50 µm.

**Figure 2.**
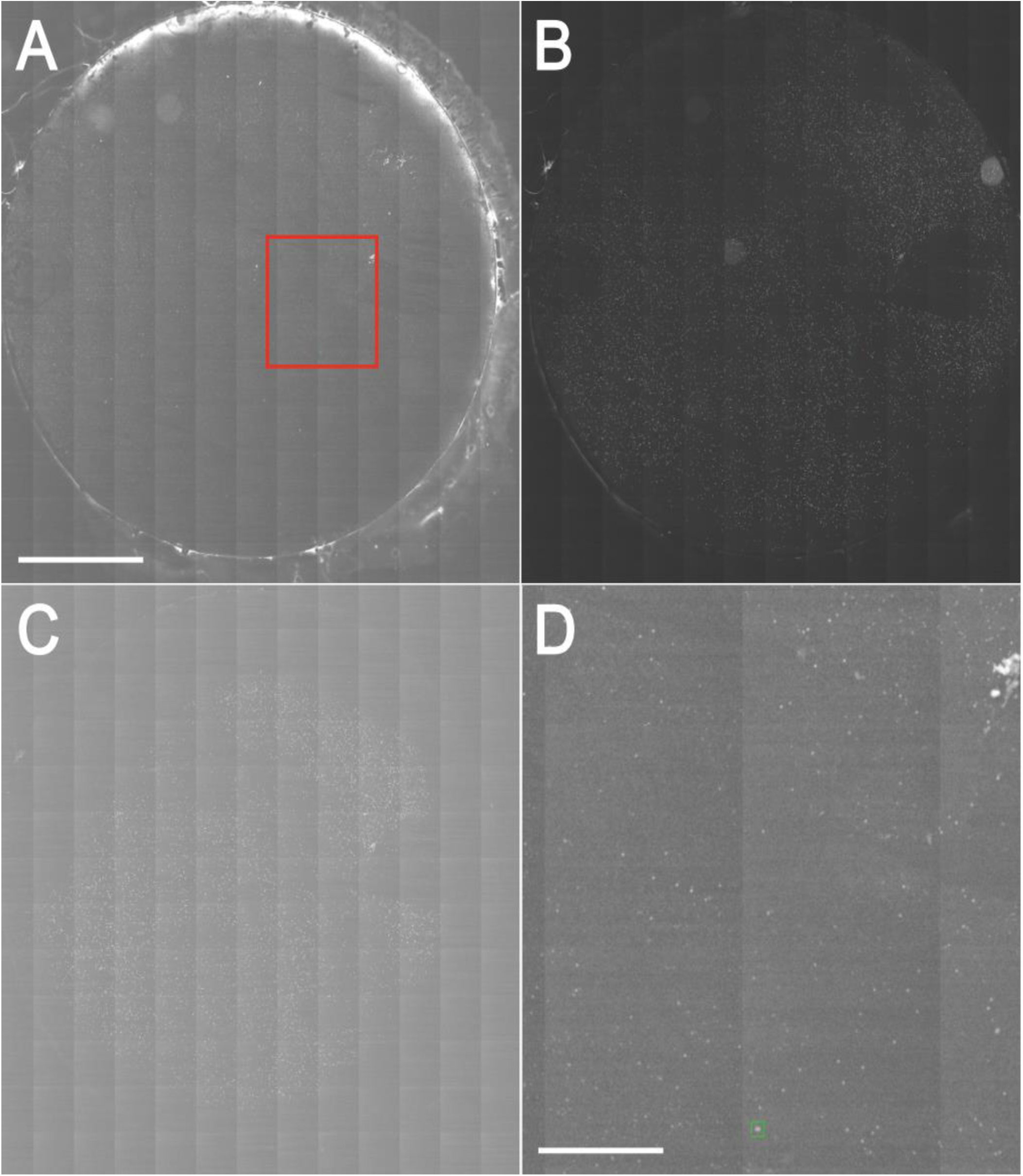
CTC detection in coverslip multiplex microscopy images. The coverslip is imaged with 169 tile images (13 x 13). Magnification, 10x. (A) Raw image of PSMA labeling in CTCs. Green square marks the detected CTC. Red square marks the area shown in (D). Scale bar equals 2 mm. (B) DAPI nuclear staining. (C) CD45 leukocytes labeling. (D) Zoom-in of the image in (A). Green square marks the detected CTC. Scale bar equals 700 µm. See Fig. S4 for another coverslip example with a CTC detection.

Our investigation demonstrated that in PSMA datasets imaged with high (63x) magnification, additional markers are not required to segment the areas with PSMA labeling. The raw PSMA imaging data, shown in Fig. 3A, often contains clusters of cells in high proximity (see the encircled area for an example of a cluster with four cells). That presented us with two problems - to (i) correctly identify that there are four cells in the cluster and (ii) precisely segment the cellular areas (or boundaries) of each cell. To this end, we first processed the raw image with a stationary wavelet transform (Olivo-Marin, 2002) and this step identified clusters of pixels with high intensity values (Starck et al., 2000), which we called “seeds” (Jones et al., 2005). In the same region highlighted with a red circle on Fig. 3B, we detected four bright pixel clusters. As our next step, we aimed to outline the precise contours based on the detected seeds and the areas were refined using an active contour algorithm (Caselles et al., 1997; Chan and Vese, 2001). This step, however, fused three of the areas, as seen within the circle in Fig. 3C. We then applied a watershed transformation (Meyer, 1994) based on the seeds shown in Fig. 3B. The watershed results are presented in Fig. 3D and we can see the topological lines representing the borders of the four cellular areas mentioned above (highlighted with a red circle).

**Figure 3.**
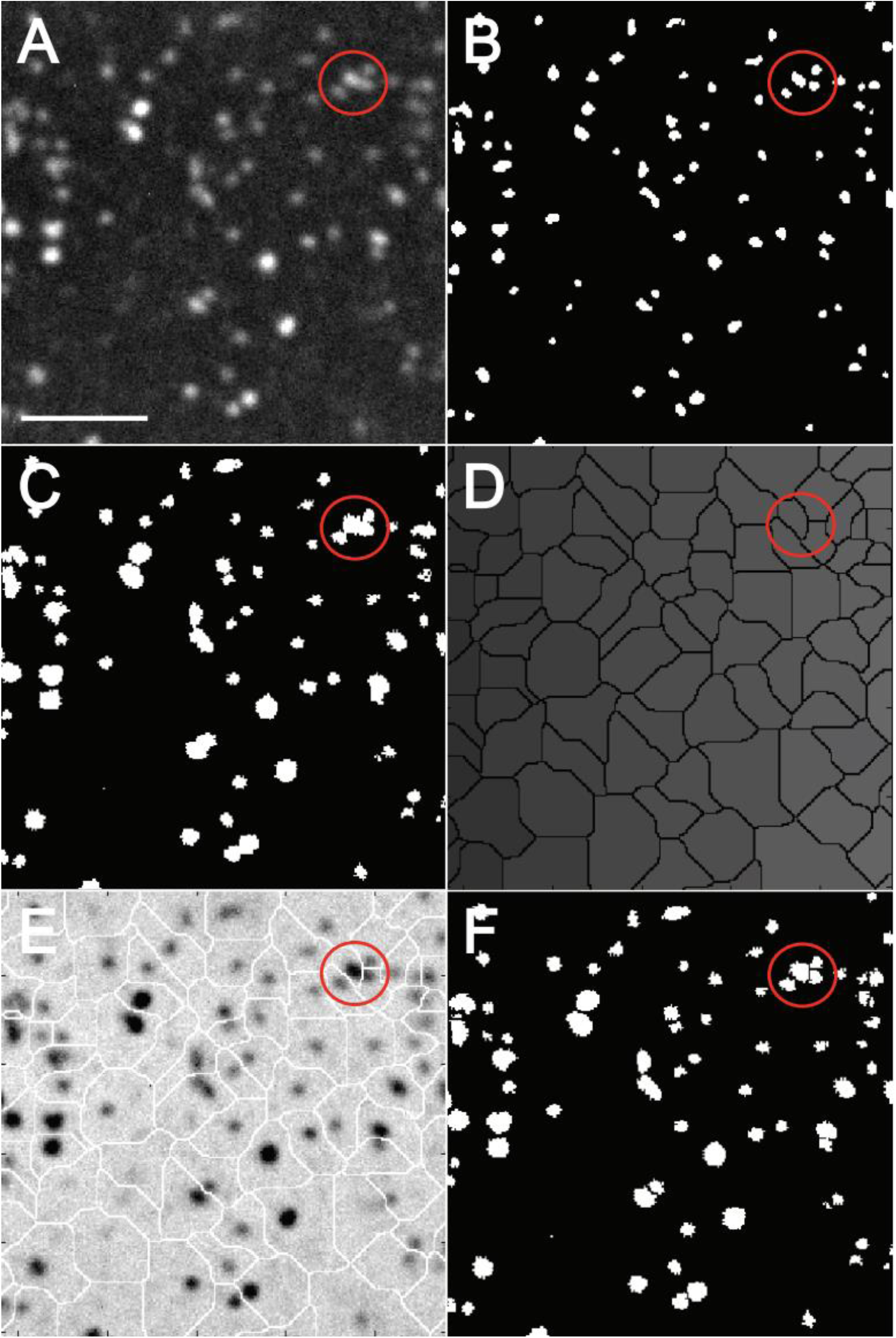
Strategy for the analysis of CTC morphology. Magnification, 63x. Scale bar equals 100 µm. (A) Raw image of PSMA labeling in CTCs. The red circle shows four cells in high proximity. (B) The initial image segmentation is accomplished by stationary wavelet transform, which identifies bright clusters in noisy images; we use this step as “seeding”. (C) Active contour, as our next step, identifies precisely the edges of the image features based on the seeds. (D) Watershed of the seeding step in (B). (E) Overlay of (C) and (D), reversed intensities. (F) Logical “and” of (C) and (D) identifies the area and their exact borders. See Fig. S5 and Fig. S6 for additional segmentation examples.

For visualization purposes and to appreciate the segmentation result, we show in Fig. 3E the overlay of Fig. 3C and Fig. 3D, both with reversed intensities, and the reader can appreciate the accuracy of the separation between the four cells within the red circle. In order to obtain the final segmentation, we performed a logical conjunction (a logical “and” operation) between the image shown in Fig. 3C and the black borders shown in Fig. 3D. This operation allowed us to improve the precision of the contours from Fig. 3C by introducing pixels with zero intensity (Fig. 3D) as borders between any fused cellular areas. The improved segmentation of the four discussed cells and can be seen in the red circle in Fig. 3F. It depicts a correct representation of the segmentation masks of the raw image of the four cells highlighted with the red circle in Fig. 3A.

Additional two examples of PSMA segmentation based on one imaging channel only (without a need of nuclear labeling or labeling of the leukocytes) are available in Fig. S5 and Fig. S6. This way, we can precisely identify the expression levels of PSMA in patient cells collected from a peripheral blood draw before and during therapy. This method can aid in the drug selection and the evaluation of any changes in the regimen based on the *ex vivo* analysis of patient cells (Matov, 2024d; Matov, 2024f; Matov, 2024g; Matov, 2025b).

### Analysis of patient-derived organoids

Our approach to image segmentation can also be utilized in the analysis of growth kinetics of patient tumors based on the *ex vivo* analysis of patient-derived organoids. After seeding of patient cells in Matrigel, organoids form and the rates of proliferation of individual clones determines the size of the organoids (Fig. 4). To image organoids, we compiled tile images consisting of 20 images (4 x 5) in order to cover the full area of the Matrigel drop. When tumors proliferate, they form spheres within which a complete anoxia at their center occurs once they reach a diameter of about 160 µm (Thomlinson and Gray, 1955). In our organoid culture, we observed very similar behavior, as the organoids exhibited a clear necrotic core once their diameter exceeded 150 µm (Fig. S7). While metastatic castrate-resistant PC organoids form circular organoids, organoids derived from drug-naïve micro-metastatic (Fig. S8) and organ-confined PC (Fig. 5) form shapes which mimic their tissue of origin. In this context, morphology and texture analysis can deliver valuable information regarding an optimal treatment strategy.

**Figure 4.**
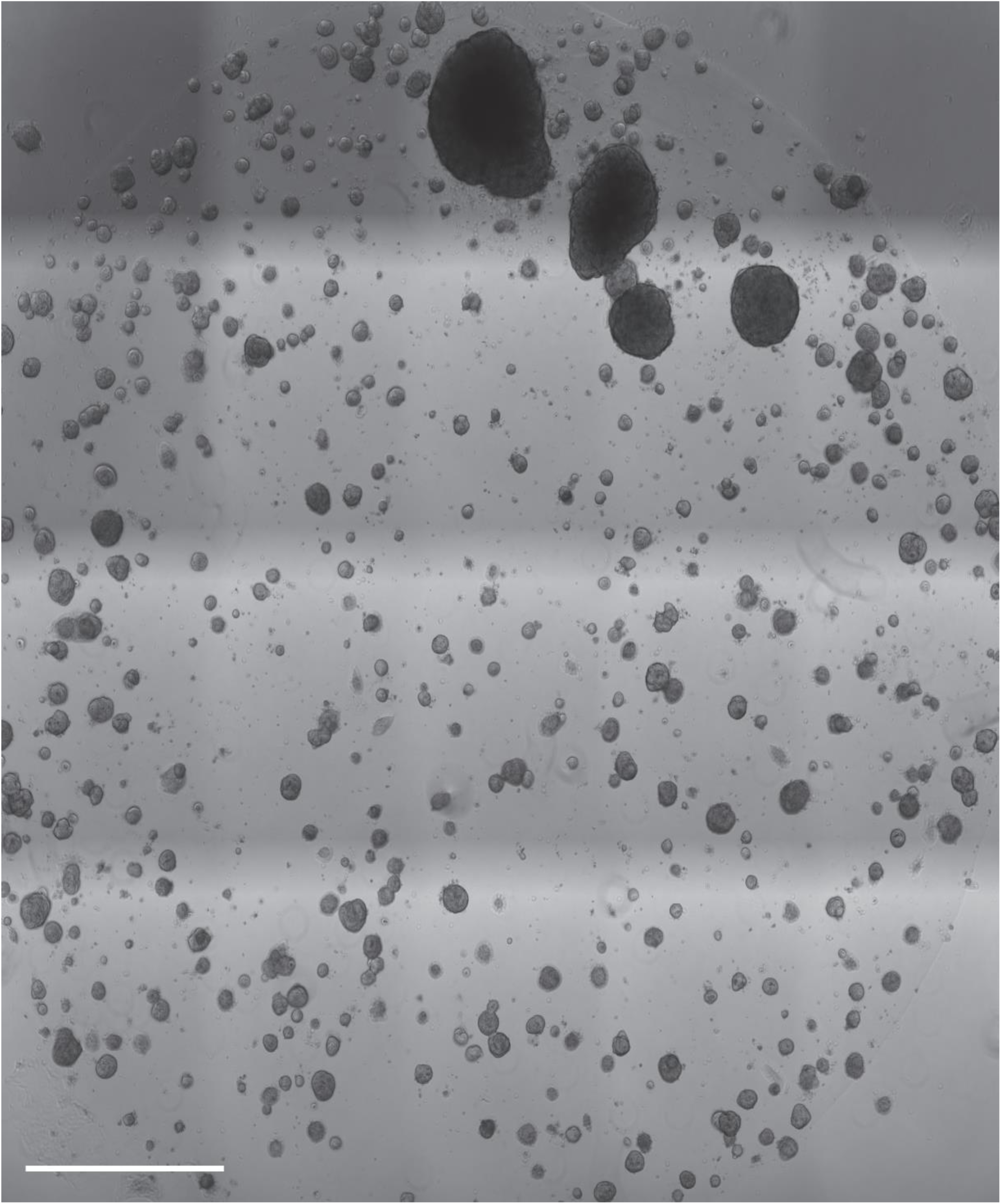
Retroperitoneal lymph node metastasis patient-derived prostate cancer organoids. Image shows a 30 µL Matrigel drop. The diameter of the Matrigel drop is 60 mm. Transmitted light microscopy, magnification 4x. Scale bar equals 12 mm. The image is composite and consists of 20 tile images (4×5). Because of the large image size (the original image is 44 MB), it is difficult to see the organoids and a compressed 22 MB image is uploaded for viewing at: https://github.com/amatov/TextureFlowAnalysis/blob/master/Large%20Image_no_drug.tif

**Figure 5.**
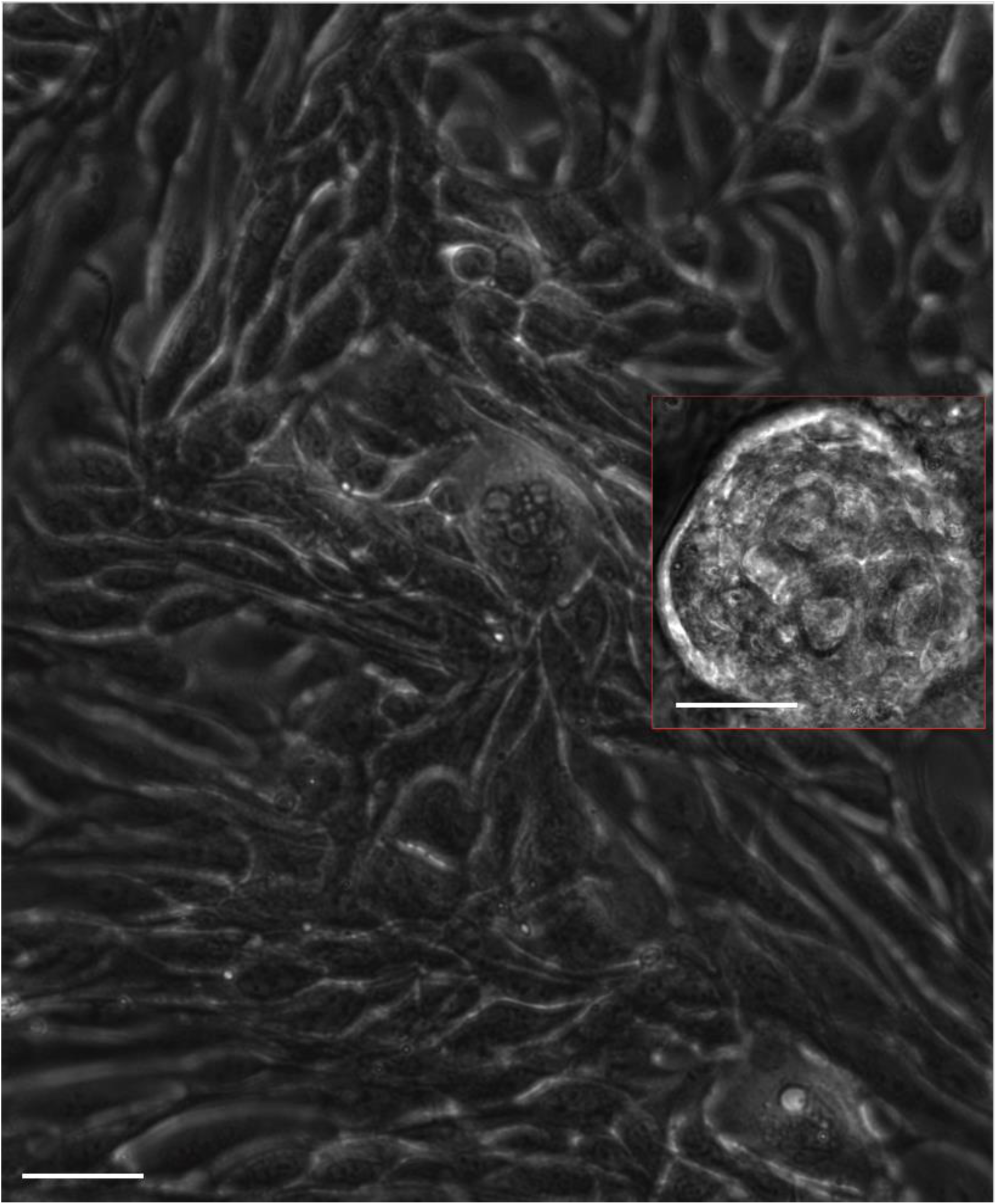
Intraductal prostate cancer organoids obtained from organ-confined disease. Organoids derived from radical prostatectomy tissue. Ductal prostate cancer is rare and very aggressive. The organoids formed visible ducts and were surrounded by a niche of fibroblasts. Organoids were often shaped like two urethral sinuses. A very large number of ductal cancer organoids tended to form at the edge of the Matrigel in direct contact with the culture medium. Phase contrast microscopy, 20x magnification. Scale bar equals 35 µm. Inset: Scale bar equals 60 µm.

When prostate tumors escape the capsule, they form micro-metastatic lesions at the retroperitoneal lymph nodes. In our organoid culture (Fig. S8), such drug-naïve tumors formed slow-proliferating tumors and had driver mutations, such as homozygous mutations in *PTEN* (exon2, c.G194>C, p.C65S) and *BRCA2* (exon14, c.T7397>C, p.V2466A).

After an initial couple of days of metabolic recovery after tissue dissociation, the tumor cells began forming organoids and about 65% of the organoids were growing – a number that decreased to about 35% within a month and remained at similar values over the six months we kept the organoids in culture (Matov, 2024a; Matov, 2025b). The organoid growth kinetics were remarkably rapid in the first about 10 days and later slowed down, reaching a plateau in about three months. After that, the organoids continued growing in size at a very slow pace. Larger organoids displayed a twined structure, suggesting a proliferative phenotype related to the mutation in *PTEN*. Many organoids formed columnar shapes of acinar adenocarcinoma, consistent with luminal differentiation (Agarwal et al., 2015).

Occasionally, the organoids appeared glandular and formed ducts – our hypothesis is that this was a result of the *BRCA2* mutations (see the large organoid in Fig. S8). The organoids did not propagate well when replated in new Matrigel and did not readily form new organoids if trypsinized to single cells. Overall, it seemed that the growth conditions were not optimal beyond the initial 10 days. Further, organoids that were kept longer in the same Matrigel after the Day 0 seeding exhibited significantly faster growth kinetics, which was likely the result of co-culture with extracellular matrix and other components of the tumor microenvironment. This behavior is indicative that the components of the medium should be adjusted based on the specific aberrations in the tumor, in particular in the context of drug-naive tumors, which are significantly less proliferative. In this context, morphology and texture measurements can facilitate the process of optimizing the growth conditions as well.

We derived organoids from organ-confined, drug-naïve intraductal PC with strong metastatic potential of the tumor. In our culture, the organoids formed visible prostatic ducts and were fast proliferating, surrounded by a niche of fibroblast cells (Fig. 5). Organoids were often shaped like the prostatic urethra. Noticeably, a very large number of ductal PC organoids tended to form at the edge of the Matrigel, in direct contact with the culture medium (Fig. S9).

We also derived organoids from high-grade (Gleason Score 4+3) acinar adenocarcinoma of the prostate, which readily formed visible glandular structures (Fig. S10, Vid. 1), but exhibited very low proliferation rates. Histologically, PC is defined by basal cell loss and malignant luminal cell expansion. Nevertheless, a paracrine growth factor can initiate tumorigenesis in basal cells as well (Lawson et al., 2010). Epithelial AR can function either as a tumor promoter or a tumor suppressor, and its function in prostate tumorigenesis depends on the cancer initiating signal (Memarzadeh et al., 2011). This function is particularly important in culturing prostate tumors as organoids. In our experience, paracrine signaling is key to organoid growth. Cultures that lose the cancer-associated fibroblasts, seize growth and become quiescent. That result suggests the need to optimize the culture medium and Matrigel formulation (Matov, 2025a).

We derived organoids from a 70 mg chest-wall resection of a patient with metastatic rectal cancer. When the organoids exceeded a size of 150 µm in diameter, they began forming intestinal glands or crypts (Lieberkühn, 1782). The intestinal epithelium is an integral component of innate immunity. The intestinal epithelial cells shape multitubular invaginations that form crypts, which increase the absorption surface of the tissue. At the base of the crypts, the intestinal stem cell niche enables the constant regeneration of the intestinal lining (e.g., enterocytes, endocrines, or goblet cells). These cells can proliferate, differentiate and move upwards, where they can be replaced after a couple of days (Gehart and Clevers, 2019). In our organoid culture, patient cancer cells readily formed organoids larger than ½ mm in diameter with some of the crypts exceeding 100 µm in diameter (Matov, 2024b). Treatment with 100 nM docetaxel induces changes in the crypts (Fig. 6), which can be quantified with our methodology and inform therapy. After a two week treatment, we measured significant ATP levels (Pant, 2024) and a clear residual disease. This way, we derived, for the first time, quiescent “persister” (Sharma et al., 2010) cancer cells in 3D patient-derived organoids cultured in 30 µL Matrigel drops (van de Wetering et al., 2015), that is, in their natural 3D environment (Fig. 6) and not as single patient cells cultured on a flat plastic surface in a 2D Petri dish. During drug holiday, in a reversible process, the drug-tolerant persister cells resume proliferating, form new organoids and become susceptible to drug action (Matov, 2025b).

**Figure 6.**
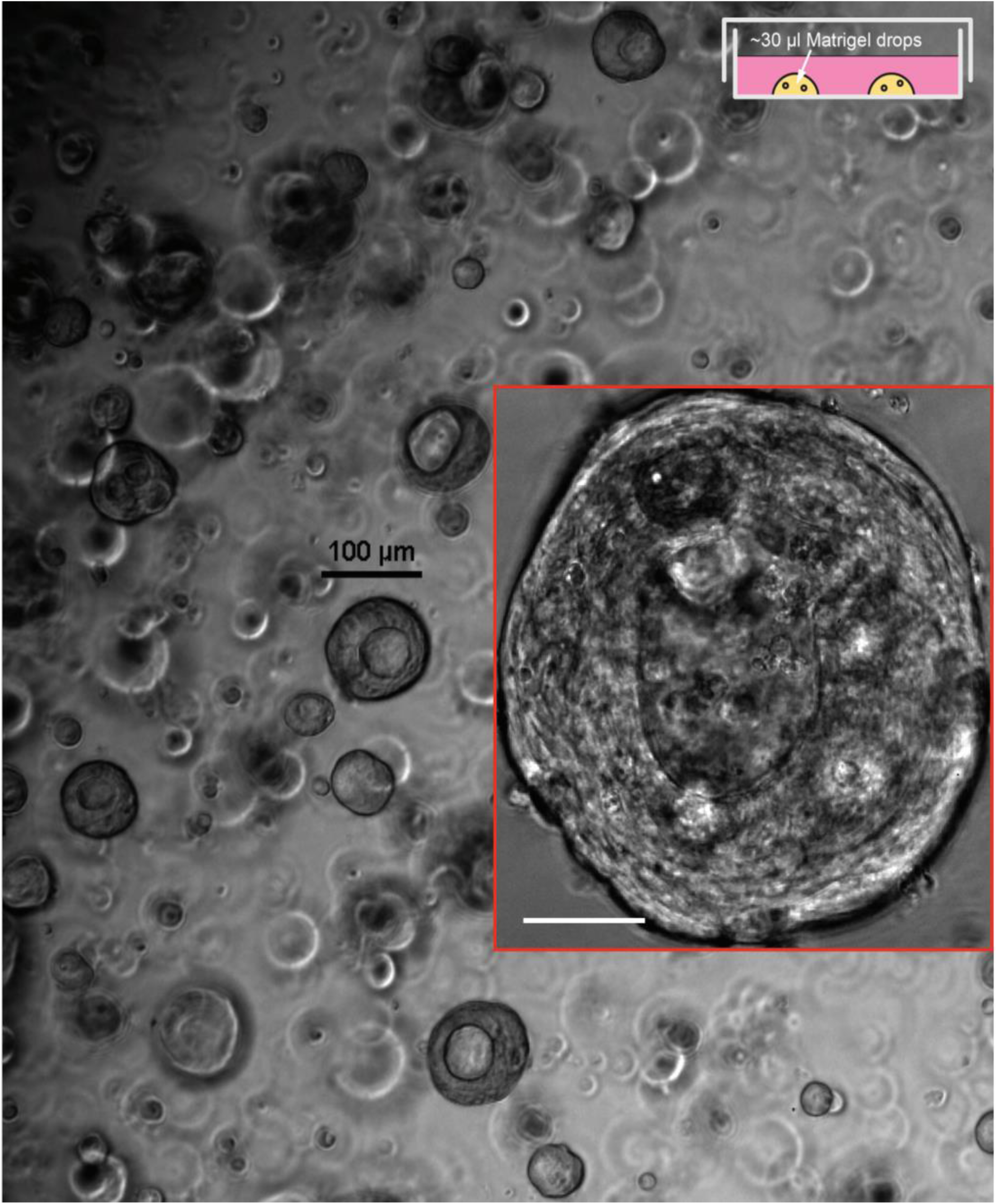
Metastatic rectal cancer organoids obtained from SU2C (chest wall) distant metastasis. Organoids with diameter 40-80 µm (with a few larger organoids also present, see the inset to the right) were plated in 30 µL Matrigel drops in six wells per condition of a 24-well plate. Transmitted light microscopy, magnification 4x. Scale bar equals 100 µm. Inset: rectal cancer organoids treated with 100 nM docetaxel. Phase contrast microscopy, magnification 20x. Scale bar equals 60 µm.

Due to a disabled antioxidant program, persister cells are unable to tolerate lipid hydroperoxide accumulation and are susceptible to ferroptotic drugs. Inducing ferroptosis is a viable strategy for the elimination of residual disease (Hangauer et al., 2017). Real-time computer vision system (Matov, 2024h), which can reliably evaluate drug action *ex vivo* will provide an additional platform for functional testing of putative compounds that exploit vulnerabilities in cancer cells, like the susceptibility to ferroptosis-inducing small molecules (Eaton et al., 2020), to eliminate residual disease. Quantitative imaging could be displayed during tumor board meetings to allow the discussion of tentative regimens based on the supporting information provided by the microscopic evaluation of patient-derived cultures.

## DISCUSSION

About a third of the patients progressing after treatment with hormonal therapies have intrinsic resistance to systemic therapy (Petrylak et al., 2004; Sweeney et al., 2015; Tannock et al., 2004). The two-thirds which respond, suffer visible side effects from drug toxicity and rapidly acquire resistance to therapy. Overall, the treatment of advanced PC is hampered by the lack of known molecular markers for reliable targeted therapies (Gao et al., 2014; Matov, 2025a). In this context, the PSMA offers a much-needed target and clinical trials are demonstrating that patients experience low levels of toxicity and manageable side effects (Tagawa et al., 2024). In this contribution, we have proposed a method to evaluate the expression of PSMA in patient cells and inform therapy. Besides PC, the expression of PSMA is elevated in other epithelial tumors (de Galiza Barbosa et al., 2020; Lauri et al., 2022), such as tumors in the kidney (Muselaers et al., 2022), breast (Unger et al., 2022), thyroid (Piek et al., 2022), liver (Thompson et al., 2022), pancreas (Sirtl et al., 2021), lung (Osman et al., 2017), and others, which shows an applicability of the approach beyond diseases of the prostate gland.

We compared our method, based on the analysis of PSMA labeling only, to analysis of multiplex microscopy imaging and generated also datasets for which PSMA images were acquired in combination with positive epithelial cell, nuclear identifiers (DAPI staining) and negative, leukocyte cell identifiers (CD45 staining) (Galletti et al., 2014a; Galletti et al., 2014b; Gleghorn et al., 2010; Tasaki et al., 2014). However, the three-color datasets did not offer any computational benefit and did not improve on our ability to correctly identify the tumor cells and the areas with PSMA expression in 63x magnification images. This result highlights the ability of this computational clinical method to detect disease and monitor drug activity based on a peripheral blood draw, followed by a simple processing step, and a straightforward quantitative microscopy method. It also demonstrates that PSMA labeling alone is sufficient to detect several types of epithelial tumors.

The treatment of PC has been hampered by the lack of availability of disease models (Gao et al., 2014) that can offer clinically relevant biomarkers of disease progression and treatment efficacy. We propose the utilization in the clinical practice of novel biomarkers based on quantitative image analysis of protein labeling in patient-derived cells. *Ex vivo* measurements of the changes in protein morphology and localization after drug treatment in patient cells can help anticipate drug resistance and design new combination regimens. Drug-induced modulation of treatment-assessing parameters in patient-derived cultures could be utilized as a clinical tool for the selection of putative targets in refractory disease, for which currently there are no available treatment options. The precise evaluation of the changes in the regulation of tumor cells will offer personalized targets based on the identified cancer vulnerabilities. Further, the analysis methodology we propose can be useful in dissecting the mechanisms of drug response in ultimate responders and these particular mechanisms can be induced in resistant tumors to achieve a complete remission. Our hypothesis is that the sensitization of resistant tumors could be done via modulation of the cytoskeleton (Matov, 2024d; Matov, 2024e; Matov, 2024g; Matov, 2025b).

We propose the utilization of the described PSMA (and DNA fragmentation (Matov, 2024c)) analytics to generate embeddings (Vaswani et al., 2017) for a generative transformer network in which the tokens are the texture metrics of the organoids. The embeddings can include morphology metrics before and after treatment with different drugs. We will retrain a transformer network with a new set of tokens - with shape descriptors rather than words and with growth/killing curves (that is, sigmoid curves) rather than sentences of human speech, that is, we will retrain a large language model (LLM) with organoid morphology data. This will allow training of the network to classify each organoid as sensitive or resistant and further identify subclasses with different mechanisms of resistance. Further, after training a generative transformer network with organoid texture and live-cell dynamics (Matov, 2024h) datasets, we will be able to model disease progression as well.

Our objective has been to present the community with an integrated, easy to use by all, tool (Matov, 2024h) for resolving the complex cellular organization and it is our goal to have such software system approved for use in the clinical practice. A key novelty is that our computational platform outputs results in real time, during imaging and without having to store the data first, which will facilitate and significantly speed up research and clinical efforts by providing instant delivery of data analysis. Augmented reality artificial intelligence software (Chen et al., 2019) can be added to live-cell microscopes, thus providing instant visual feedback with added graphics during sample observation. This functionality will enhance the images by also displaying analysis results describing the measured differences (Matov, 2024i) and, in doing so, facilitate the image interpretation by the clinical operator. It will significantly enhance the ability of clinicians to make quick and correct decisions regarding drug action.

Real-time image analysis (Matov, 2024i) can also assist surgeons with lesion localization during surgical resections (Deivasigamani et al., 2023). The software will segment the precise area of selected fiducial biomarkers on-the-fly, enhance the images, and analyze the time evolution of their contour to improve the precision of the resection boundary. Similarly, it will improve the laser positioning guidance during multi-parametric MRI-guided focal laser ablation therapy (van Luijtelaar et al., 2019).

The changes occurring in disease are consequences of the modulation of the metabolic patterns leading to pre-clinical conditions and longitudinal samples (Matov, 2024j) can be considered forming a Markov chain. Modeling of sequential input data as a Markov process is an intuitive way to utilize LLMs because of the Markovianity of natural languages (Shannon, 1951). Given a large number of longitudinal datasets, that is, consisting of samples (for each patient) before and after the detection of neoplastic transformation, training of a generative pre-trained transformer model can deliver both classification and prediction of disease progression (prognostic analysis) based on the identification of significant changes in the image datasets.

Given the availability of high-quality fundamental research equipment for high resolution live-cell microscopy in most university hospitals, what we propose is to embed in these systems real-time augmented reality software (Matov, 2024i) for automated quantification and on-the-fly statistical analysis of intracellular behavior. Introducing the quantitative imaging method (Matov, 2024h) to the clinic will allow physicians to fine-tune, with a high level of certainty, an optimal treatment regimen for each patient.

## MATERIALS AND METHODS

### Coverslip processing

Peripheral blood previously drawn from mCRPC patients (the samples were de-identified) receiving docetaxel-based first line chemotherapy was thawed and processed using ficolling (Ficoll-Paque®) to separate cells in the buffy coat from the blood. The cells were isolated by centrifugation, fixed in phenol, and blocked on 8 mm coverslips. In brief, the ficolling method, through centrifugation, isolates peripheral blood mononuclear cells and plasma from granulocytes and erythrocytes. The mononuclear layer contains lymphocytes, monocytes, and CTCs. These cells are plated onto an 8 mm coverslip and subsequently stained for biomarkers (PSMA, DAPI, CD45, MT, and AR). PSMA labeling was done using ^177^Lu -J591, anti-PSMA monoclonal antibody J591.

### Sample processing

Small RNA were extracted from de-identified urine samples (50 mL) by Norgen Biotek Corp.

Retroperitoneal lymph node organoid whole genome sequencing (736 ng DNA) was done at the UCSF Core Facilities.

### Cell culture

Gastric cancer cell lines SCH, HS746T, MKN7, TMK1, and AZ521 were a gift from Patrick Tan; the SNU1 cells were a gift from Zev Wainberg; the MKN45 cells were a gift from Markus Möhler.

Organoids with diameter 40-80 µm were plated in 30 µL Matrigel drops in six wells per condition of a 24-well plate per condition and treated for two or more weeks with docetaxel with concentration of 10 nM, 100 nM, and 1 µM to eliminate the vast majority of the organoid cells and induce a persister state in the residual cancer cells. This treatment was followed by co-treatment with docetaxel 100 nM and small molecule ML210 5 µM or docetaxel 100 nM combined with small molecule ML210 5 µM and lipophilic antioxidants ferrostatin-1 2 µM, rescue treatment. Organoids with diameter 40-80 µm were plated in 30 µL Matrigel drops in six wells per condition of a 24-well plate per condition and also treated for two or more weeks with FOLFOX (folinic acid, fluorouracil, and oxaliplatin) (de Gramont et al., 1997) with concentration of 1 µM, 10 µM, and 100 µM to eliminate the vast majority of the organoid cells and induce a persister state in the residual cancer cells. Cell viability was read out using CellTiter Glo (Promega).

Primary and retroperitoneal lymph node metastatic prostate tumor and sternum metastatic rectal tumor tissues were dissociated to single cells using modified protocols from the Witte lab. Organoids were seeded as single cells in three 30 µL Matrigel drops in 6-well plates. Organoid medium was prepared according to modified protocols from the Clevers lab (van de Wetering et al., 2015) and the Chen lab (Gao et al., 2014).

To obtain a single cell suspension, tissues were mechanically disrupted and digested with 5 mg/mL collagenase in advanced DMEM/F12 tissue culture medium for several hours (between 2 and 12 hours, depending on the biopsy and resection performed). If this step yielded too much contamination with non-epithelial cells, for instance during processing of primary prostate tumors, the protocol incorporated additional washes and red blood cell lysis (Goldstein et al., 2011). Single cells were then counted using a hemocytometer to estimate the number of tumor cells in the sample, and seeded in growth factor-reduced Matrigel drops overlaid with PC medium (Gao et al., 2014). With radical prostatectomy specimens, we had good success with seeding three thousand cells per 30 μL Matrigel drop, but for metastatic samples organoid seeding could reliably be accomplished with significantly less cells, in the hundreds.

### Microscopy imaging

Coverslips were imaged at magnification 63x and 1.2 NA of the objective. The type of microscopy used was widefield epi-fluorescence widefield epi-fluorescence at 10x and 63x magnification with ORCA-Flash 4.0 (6.5 µm/pixel) camera.

Organoids were imaged using transmitted light microscopy at 4x magnification and phase contrast microscopy at 20x magnification on a Nikon Eclipse Ti system with camera Photometrics CoolSnap HQ2.

Organoid long-term imaging was done over 72 hours by taking an image every 10 minutes at 20x magnification.

We imaged EB1ΔC-2xEGFP-expressing organoids (Koo et al., 2013) by time-lapse spinning disk confocal microscopy using a long working distance 60x magnification water immersion 1.45 NA objective. We acquired images at half a second intervals for a minute to collect datasets without photobleaching (Gierke and Wittmann, 2012).

### Image analysis

All 10x and 63x image analysis programs for PSMA segmentation and graphical representation of the results were developed in Matlab and C/C++. The wavelet transform method used, spotDetector, was described and validated in (Olivo-Marin, 2002). The computer code is available for download at: https://github.com/amatov/SegmentationBiomarkerCTC.

The 10x image analysis programs for PSMA, α-tubulin, DAPI, and CD45 segmentation as well as 3D AR and DAPI segmentation, and graphical representation of the results were developed in Matlab and C/C++. The wavelet transform method used, spotDetector, was described and validated in (Olivo-Marin, 2002) and the unimodal pixel intensity thresholding in (Rosin, 2001). We identified the CTC areas as connected-component labeling pixel lists in the epithelial tumor imaging channel (such as PSMA or occasionally CK) for which in the nuclear imaging channel there is a DAPI stain with a statistically representative size and a circular shape. At the same time, we required that these cells are negative in the CD45 channel, that is, the absence of a leukocyte marker. Similarly, to detect neutrophils and lymphocytes, we identified clusters of bright pixels in the CD45 channel for which a nuclear area is detected in the DAPI channel and the epithelial stain is not present. On multiple occasions, we detected double-positive (PSMA+/DAPI+/CD45+ or CK+/DAPI+/CD45+) and double-negative (PSMA-/DAPI+/CD45- or CK-/DAPI+/CD45-) cells, which we classified in separate bins.

Drug-induced MT bundles were evaluated in terms of thickness and texture parameters in 63x images. The detection of tyrosinated tubulin in 10x images was considered to be a marker of drug sensitivity. We analyzed the cellular localization of AR in 3D image stacks, in which we performed segmentation in 35 individual 2D images and reconstructed the volume, and classified the CTCs as sensitive to drug treatment, if the AR is within the volume of the nucleus, or resistant, if the AR is mainly in the cytoplasm.

To calculate cell edge intensity in immunofluorescence images of drug-treated cells growing in clusters, we identify the centroid position of the nucleus by applying unimodal pixel intensity thresholding (Rosin, 2001) in the DAPI channel, which allows the segmentation of the nuclei (Fig. S11). We then used the segmented nuclear areas and dilated the images (Minkowsk, 1901). By removing from the dilated image the raw image with the nuclei, we obtained lists of pixels that form rings around each nucleus. We averaged the pixel intensities in the MT channel for each ring (Fig. S12) before and after treatment of lung cancer cells with 100 nM docetaxel. Our analysis provides a quantitative readout of drug susceptibility and resistance, as the pixel intensities have high values in drug-sensitive cells, because of the drug-induced MT bundling (Matov, 2024d), and low values in drug-resistant cells. The computer code can be utilized also for the analysis of CTCs and patient-derived organoids (Fig. S12); it is available for download at: https://github.com/amatov/SegmentationBiomarkerCTC, https://github.com/amatov/ResistanceBiomarkerAnalysis, https://github.com/amatov/AntibodyTextureMorphology.

## Data Availability

All data produced in the present study are available upon reasonable request to the authors

https://github.com/amatov/SegmentationBiomarkerCTC

## Abbreviations

AR: androgen receptor
BRCA1/2: breast cancer type 1/2 susceptibility protein
CD45: protein tyrosine phosphatase receptor type C
CK: cytokeratin
CTC: circulating tumor cell
GC: gastric cancer
DAPI: 4′,6-diamidino-2-phenylindole
EB1: microtubule end binding protein 1
EB1ΔC: EB1 construct truncated at amino acid 248 that does not interact with other proteins at the microtubule end
EGFP: enhanced green fluorescent protein (2xEGFP indicates two molecules)
FOLFOX: folinic acid, fluorouracil, and oxaliplatin
GPX4: glutathione peroxidase 4
MT: microtubule
PARP: poly (ADP-ribose) polymerase (ADP – adenosine diphosphate)
PC: prostate cancer (mCRPC – metastatic castrate-resistant prostate cancer)
PSMA: prostate-specific membrane antigen

## DECLARATIONS

### Ethics approval and consent to participate

IRB (IRCM-2019-201, IRB DS-NA-001) of the Institute of Regenerative and Cellular Medicine. Ethical approval was given.

Approval of tissue requests #14-04 and #16-05 to the UCSF Cancer Center Tissue Core and the Genitourinary Oncology Program was given.

Informed consent to participate was obtained for clinical studies with IRB protocols 0804009740 and 0707009283.

Informed consent was obtained from all subjects and/or their legal guardian(s). The study adhered to the Declaration of Helsinki.

### Consent for publication

Consent for publication was given.

### Availability of data and materials

The datasets used and/or analyzed during the current study are available from the corresponding author upon reasonable request. The prostate cancer organoid whole genome sequencing datasets (https://www.ncbi.nlm.nih.gov/bioproject/1255230) are accessible at: https://www.ncbi.nlm.nih.gov/sra/SRX28551163.

### Authors’ contributions

A.M. conceived the study, cultured patient-derived organoids and cancer cell lines, performed high resolution microscopy imaging, designed the analysis algorithms, wrote the proof-of-principle code to demonstrate the feasibility of segmentation of PSMA, DAPI, CD45, MT, and AR markers via stationary wavelet transform and unimodal pixel intensity thresholding, wrote code for the segmentation of MT bundles and AR nuclear localization in 10x and 63x images, and prepared the manuscript.

## Acknowledgements

I thank Shayan Modiri for adding the watershed transformation and active contour for PSMA analysis in 63x images. I also thank Guang-Yu Lee for processing coverslips and acquiring CTC images, and Nikolay Mihaylov for running the code on tile 10x image datasets. Giuseppe Galletti performed the IC50 experiments. The patient blood samples analyzed were from clinical studies with IRB protocols 0804009740 and 0707009283 at Cornell Medicine. I am grateful to the Genitourinary Tissue Utilization committee and the Genitourinary and Prostate SPORE Tissue Cores at the UCSF Cancer Center for the approval of my tissue requests #14-04 and #16-05, the Stand Up To Cancer / Prostate Cancer Foundation (SU2C/PCF) West Coast Dream Team (WCDT), the Institute of Regenerative and Cellular Medicine for issuing the Institutional Review Board protocol approval IRCM-2019-201, IRB DS-NA-001 for the observational study “Longitudinal analysis of next-generation sequencing of nucleic acids for early detection of degenerative diseases such as cardiovascular, neoplastic and diseases related to the nervous system”, and James Faber for his feedback regarding the protocol and the process of approval.

## SUPPLEMENTARY MATERIALS

**Figure S1.**
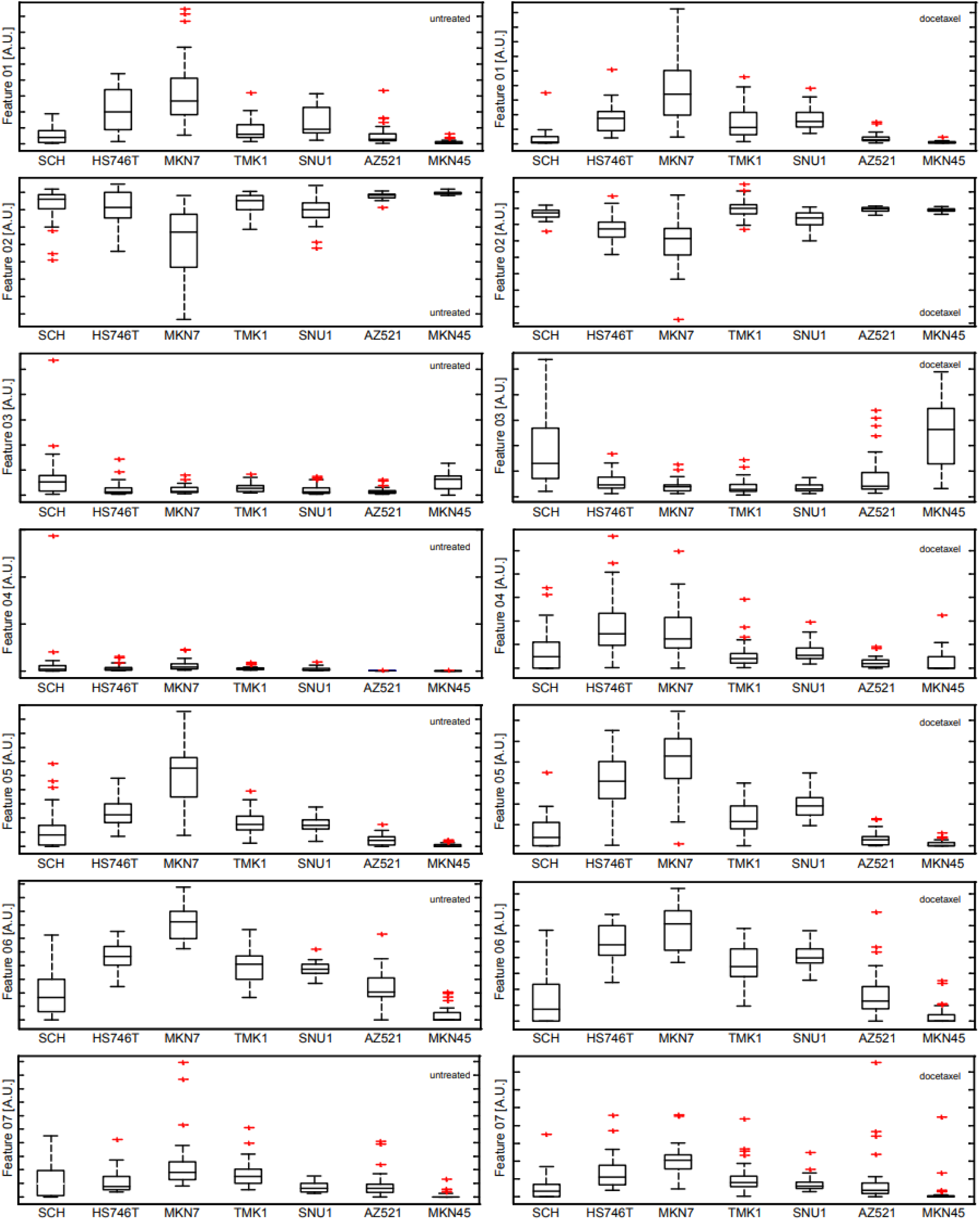
Analysis of changes in MT network morphology and localization. The distributions of features 1-7 (of 21) are shown for populations of 50 cells for 7 diffuse gastric cancer cell lines before (left side) and after (right side) treatment with 100 nM docetaxel.

**Figure S2.**
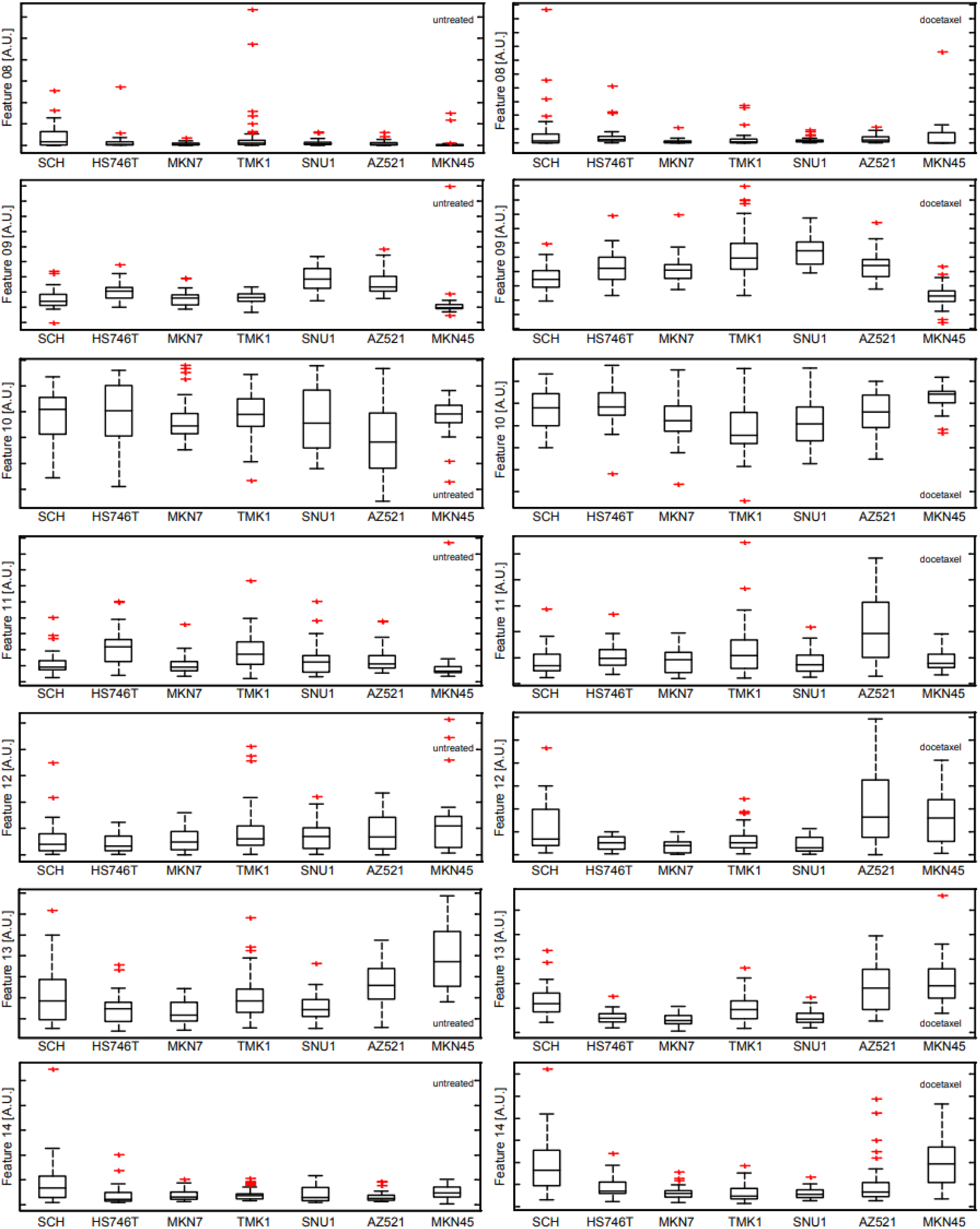
Analysis of changes in MT network morphology and localization. The distributions of features 8-14 (of 21)

**Figure S3.**
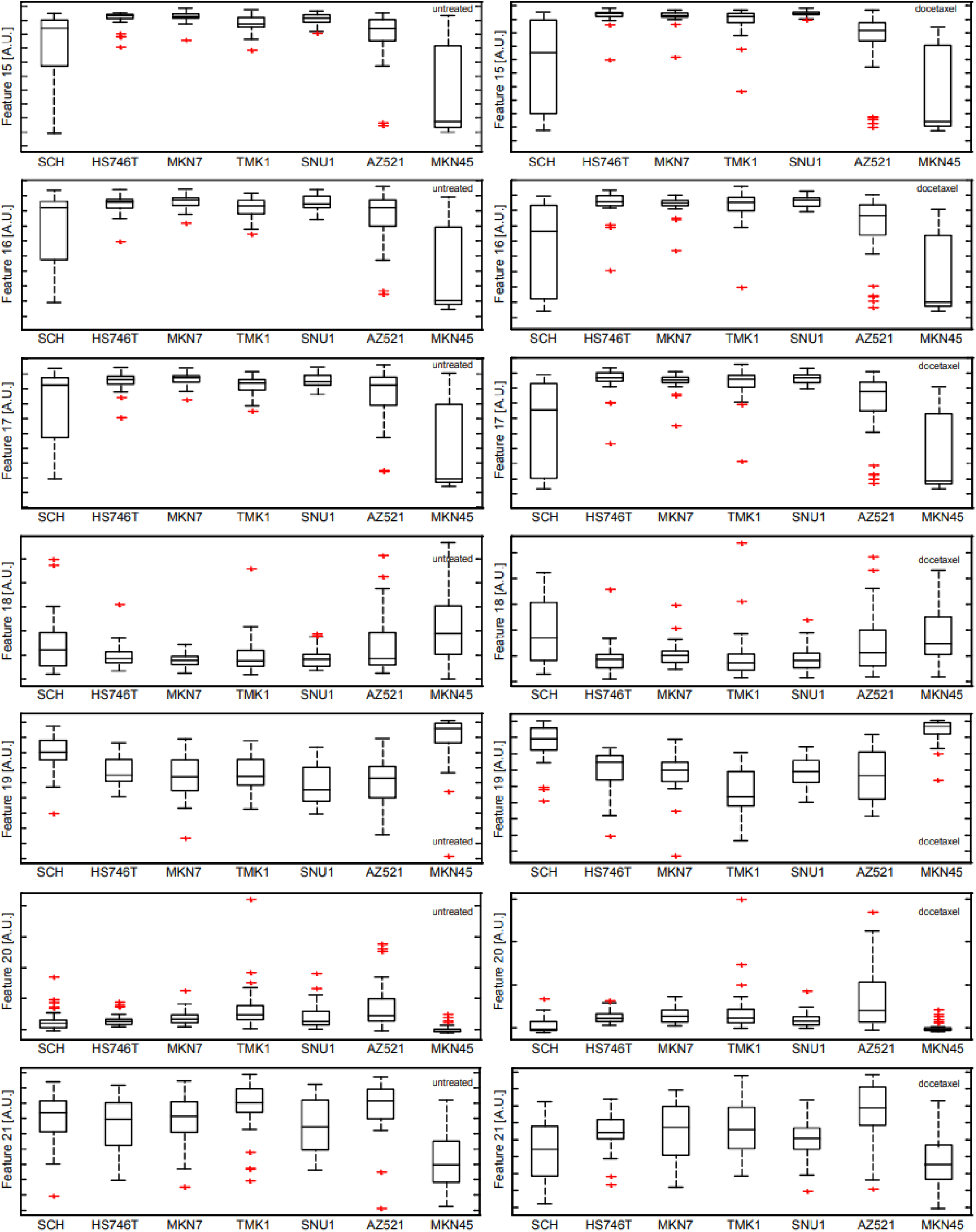
Analysis of changes in MT network morphology and localization. The distributions of features 15-21 (of 21)

**Figure S4.**
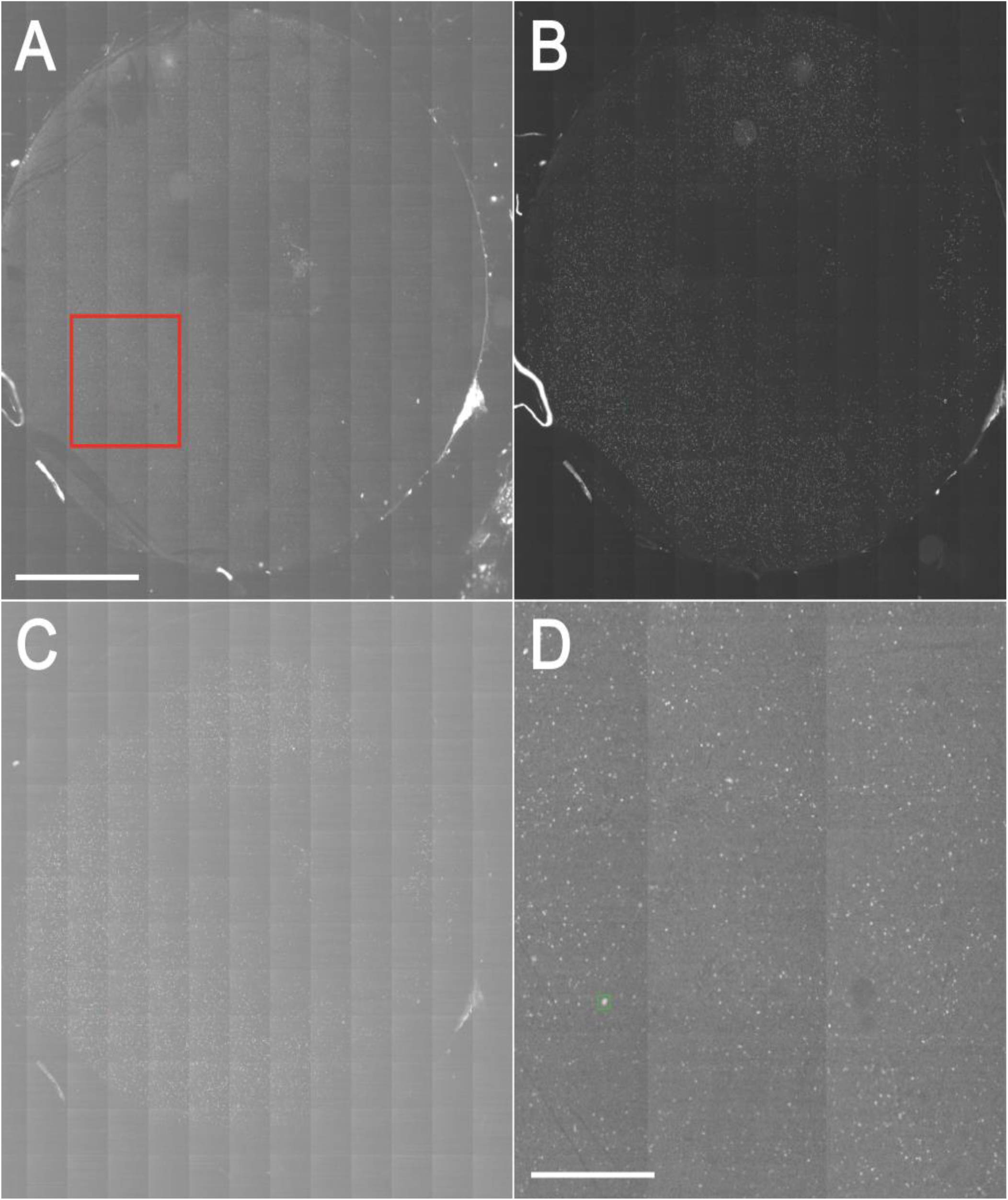
CTC detection in coverslip multiplex microscopy images. The coverslip is imaged with 169 tile images (13 x 13). Magnification, 10x. (A) Raw image of PSMA labeling in CTCs. Green square marks the detected CTC. Red square marks the area shown in (D). Scale bar equals 2 mm. (B) DAPI nuclear labeling. (C) CD45 leukocytes labeling. (D) Zoom-in of the image in (A). Green square marks the detected CTC. Scale bar equals 700 µm.

**Figure S5.**
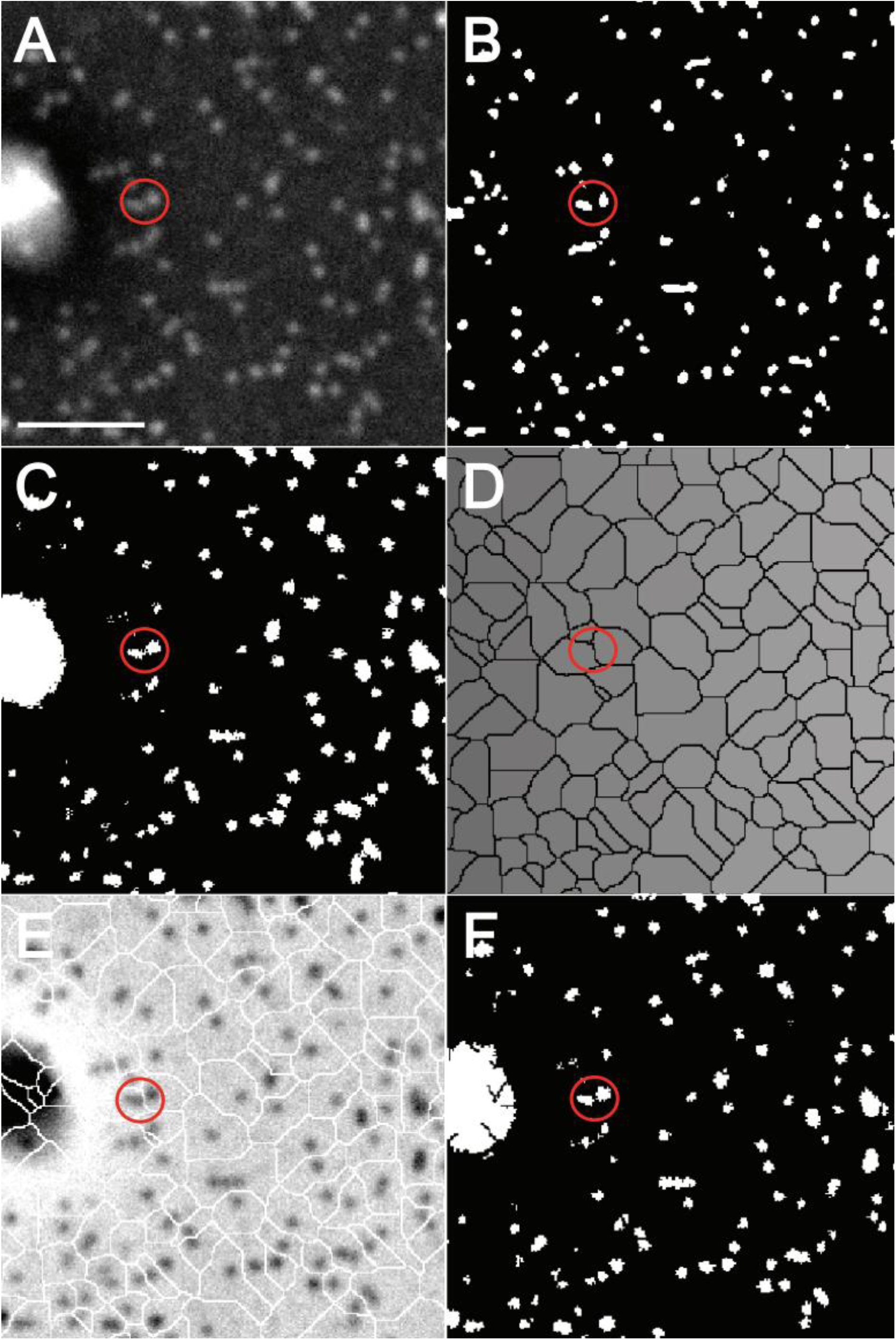
Strategy for the analysis of CTC morphology. Magnification, 63x. Scale bar equals 100 µm. (A) Raw image of PSMA labeling in CTCs. The red circle shows two cells in high proximity. (B) The initial image segmentation is accomplished by stationary wavelet transform, which identifies bright clusters in noisy images; we use this step as “seeding”. (C) Active contour, as our next step, identifies precisely the edges of the image features based on the seeds. (D) Watershed of the seeding step in (B). (E) Overlay of (C) and (D), reversed intensities. (F) Logical “and” of (C) and (D) identifies the area and their exact borders.

**Figure S6.**
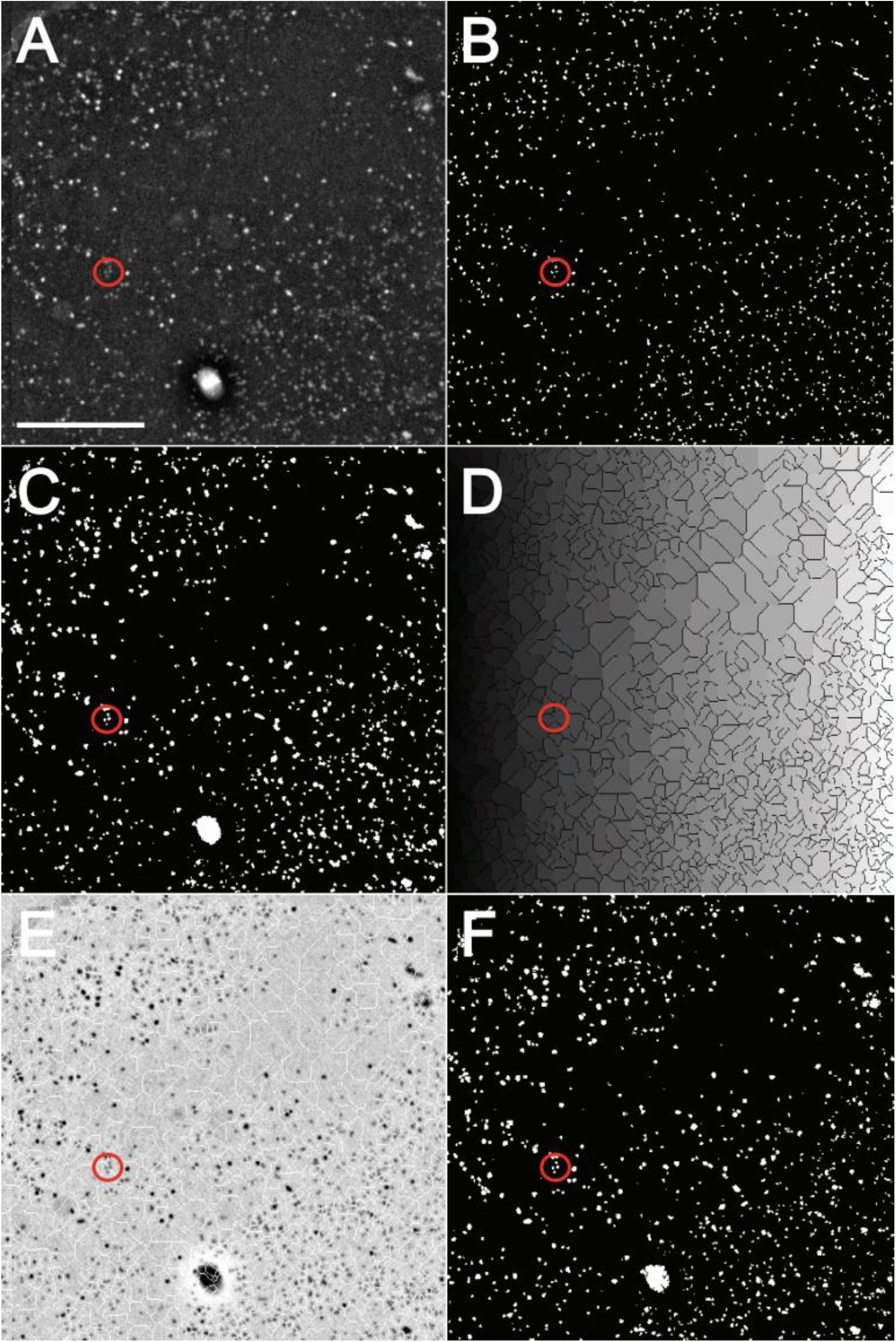
Strategy for the analysis of CTC morphology. Magnification, 63x. Scale bar equals 200 µm. (A) Raw image of PSMA labeling in CTCs. The red circle shows four cells in high proximity. (B) The initial image segmentation is accomplished by stationary wavelet transform, which identifies bright clusters in noisy images; we use this step as “seeding”. (C) Active contour, as our next step, identifies precisely the edges of the image features based on the seeds. (D) Watershed of the seeding step in (B). (E) Overlay of (C) and (D), reversed intensities. (F) Logical “and” of (C) and (D) identifies the area and their exact borders.

**Figure S7.**
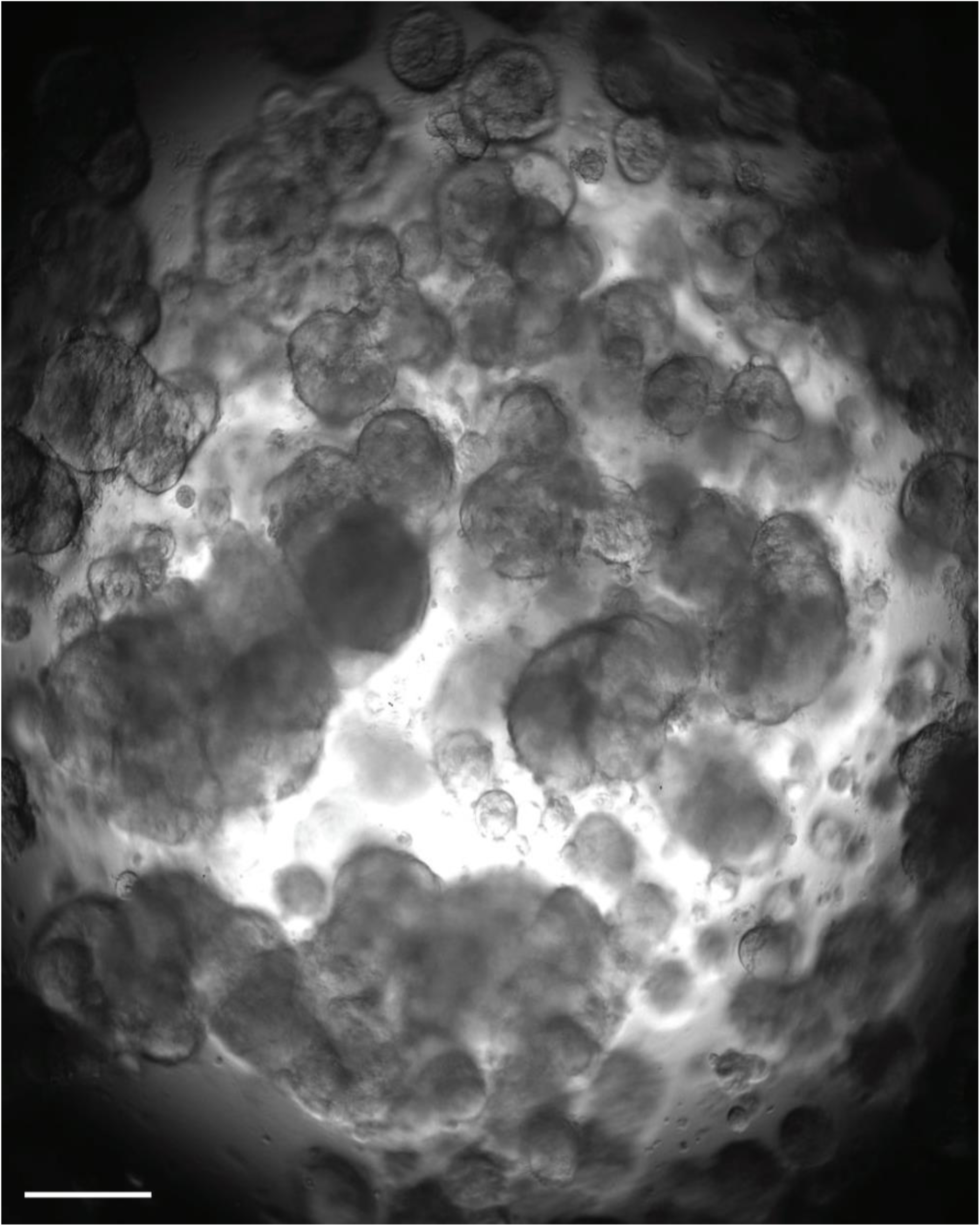
Retroperitoneal lymph node metastasis patient-derived prostate cancer organoids. Organoids with diameter 40-80 µm were plated in 30 µL Matrigel drops and cultured for five weeks. Transmitted light microscopy, 4x magnification. Scale bar equals 200 µm.

**Figure S8.**
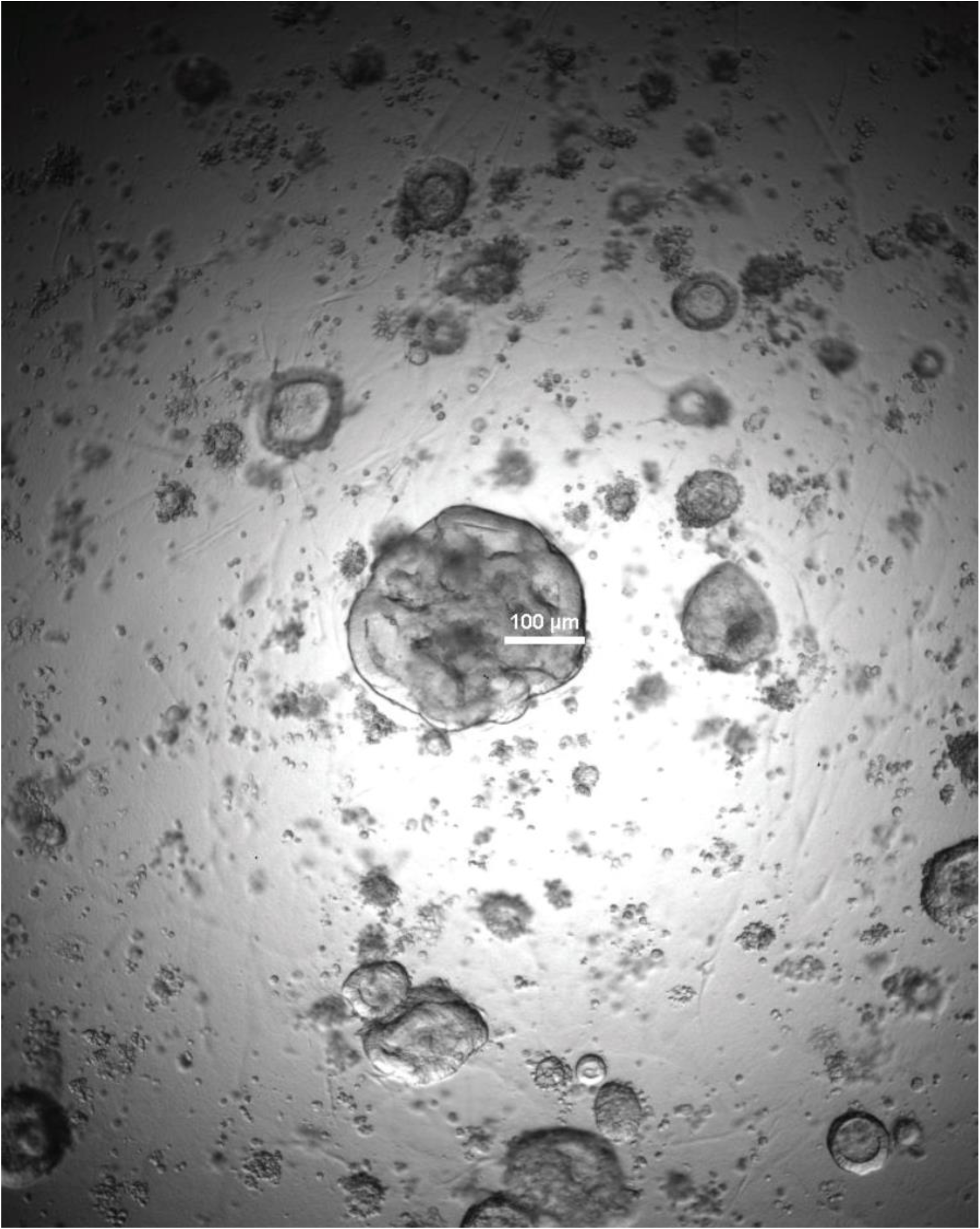
Metastatic prostate cancer organoids obtained from micro-metastasis at a retroperitoneal lymph node. Organoids derived from radical prostatectomy tissue. Day 105. Organoid whole genome sequencing showed homozygous mutations in *PTEN* and *BRCA2*. Transmitted light microscopy, 4x magnification. Scale bar equals 100 µm.

**Figure S9.**
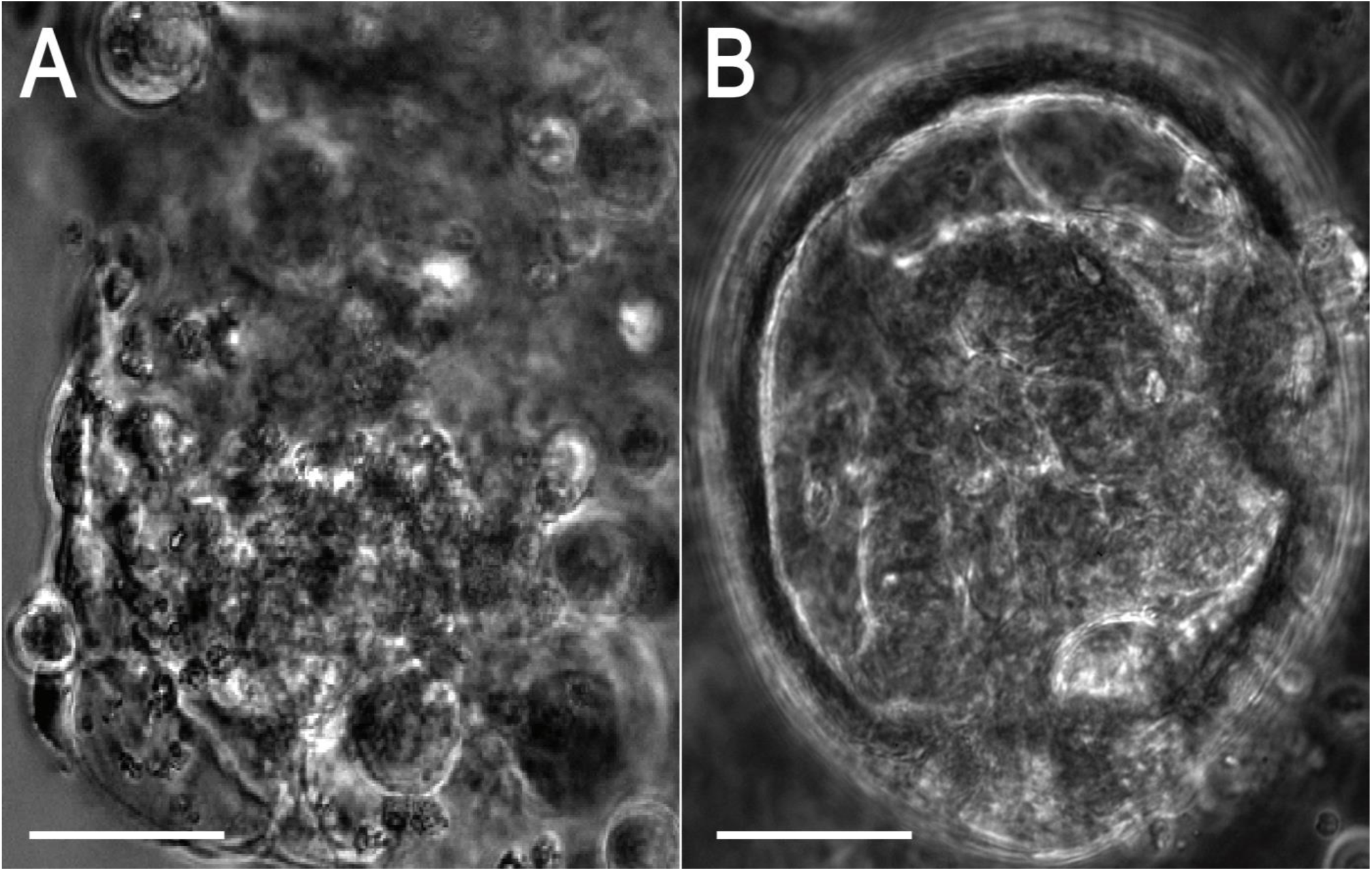
Intraductal prostate cancer organoids obtained from organ-confined disease. Organoids derived from radical prostatectomy tissue. Ductal prostate cancer is rare and very aggressive. The organoids formed visible glandular structures. Phase contrast microscopy, 20x magnification. (A) Ductal cancer organoids tended to form at the edge of the Matrigel in direct contact with the culture medium. Scale bar equals 50 µm. (B) Organoids shaped as the prostatic urethra. Scale bar equals 60 µm.

**Figure S10.**
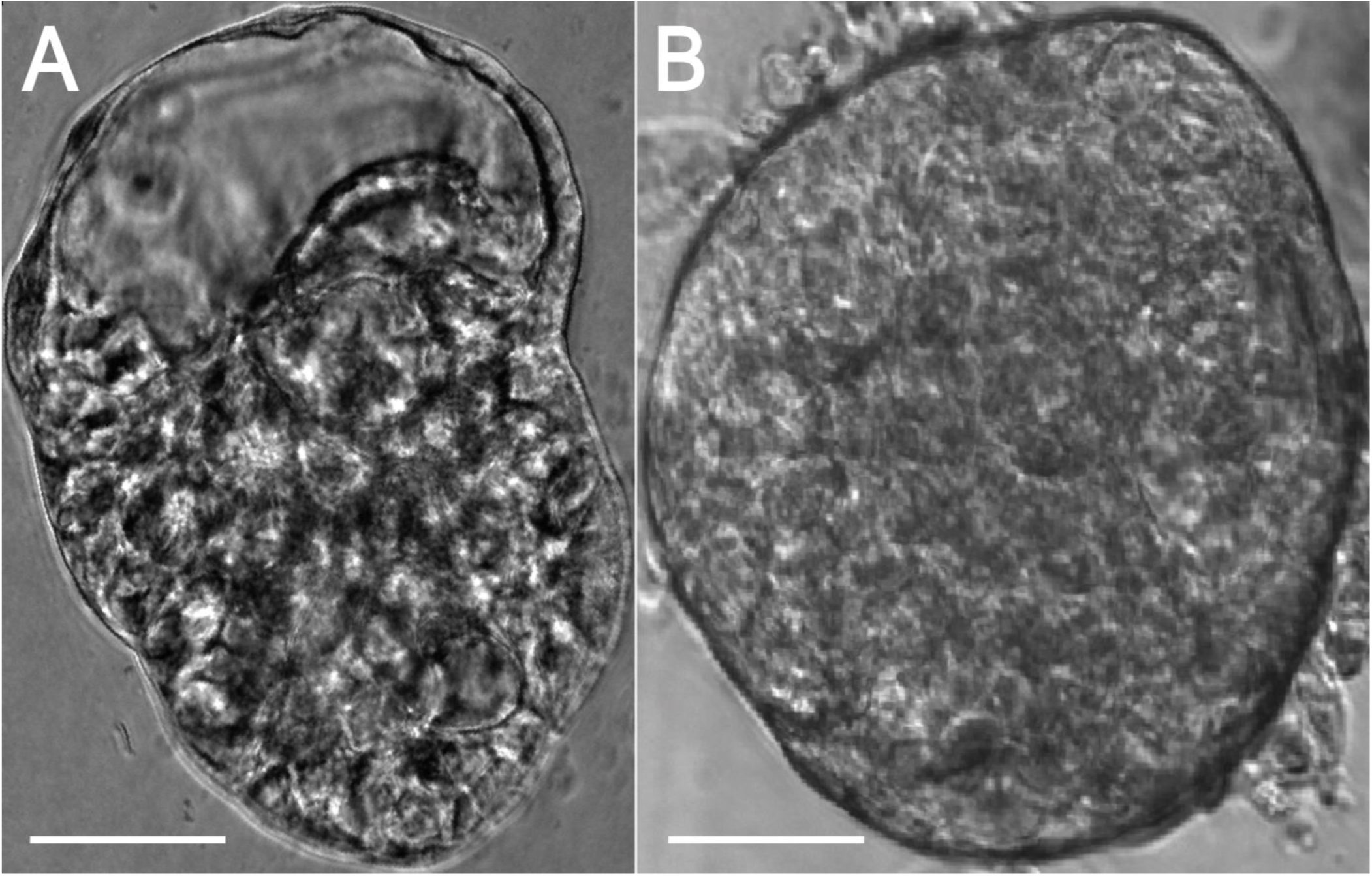
Acinar adenocarcinoma organoids obtained from organ-confined disease. Organoids derived from radical prostatectomy tissue. The organoids formed visible glandular structures. Phase contrast microscopy, 20x magnification. (A) The organoids tended to form glands (see also Vid. 1). Scale bar equals 70 µm. (B) Organoid cells formed acini. Scale bar equals 60 µm.

**Figure S11.**
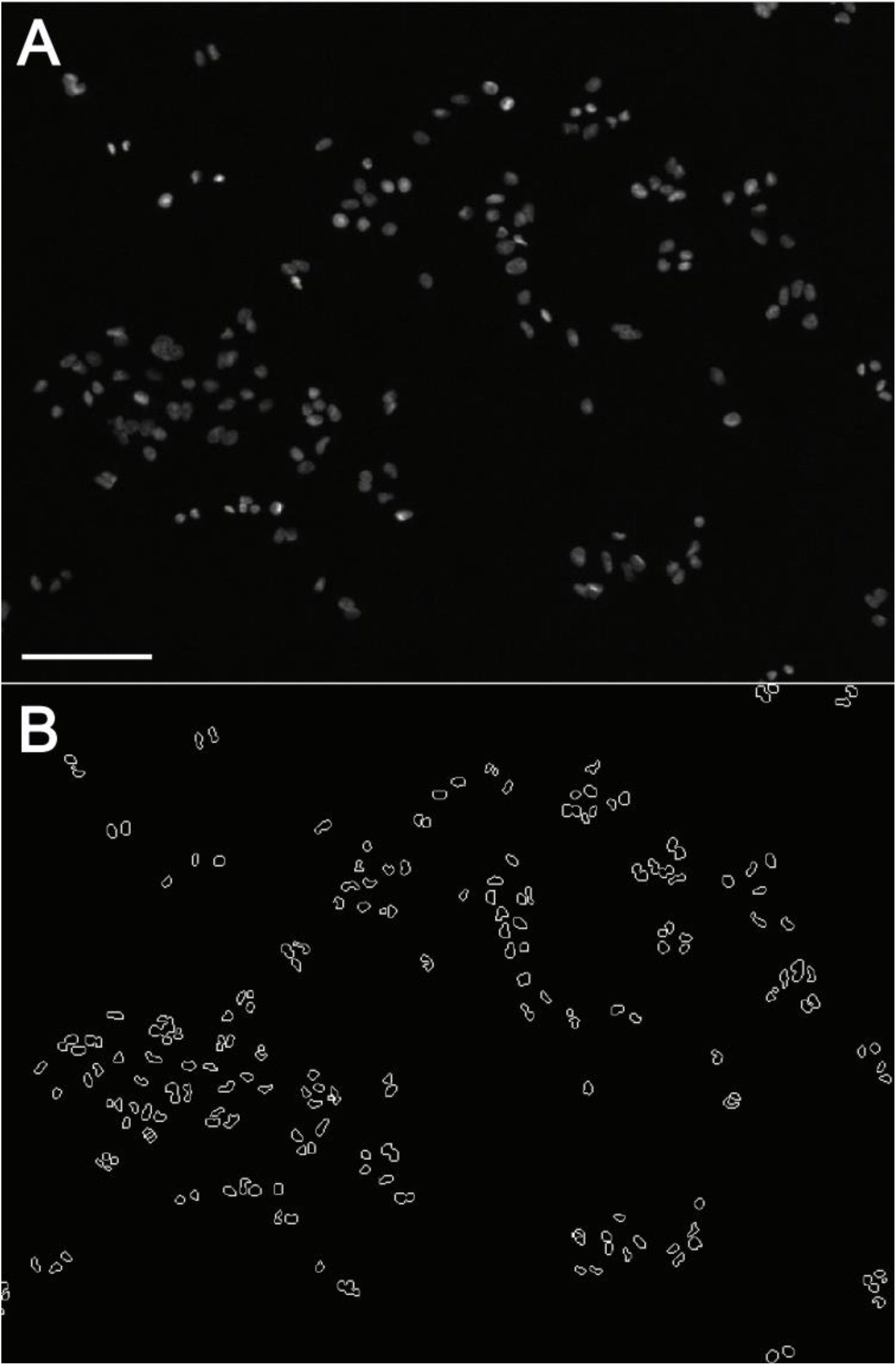
Segmentation of nuclear areas of A549 lung cancer cells. DAPI is fluorescently labeled. The figure presents an image segmentation approach to determine the outlines of the nuclei of cells growing in clusters. Scale bar equals 40 µm.

**Figure S12.**
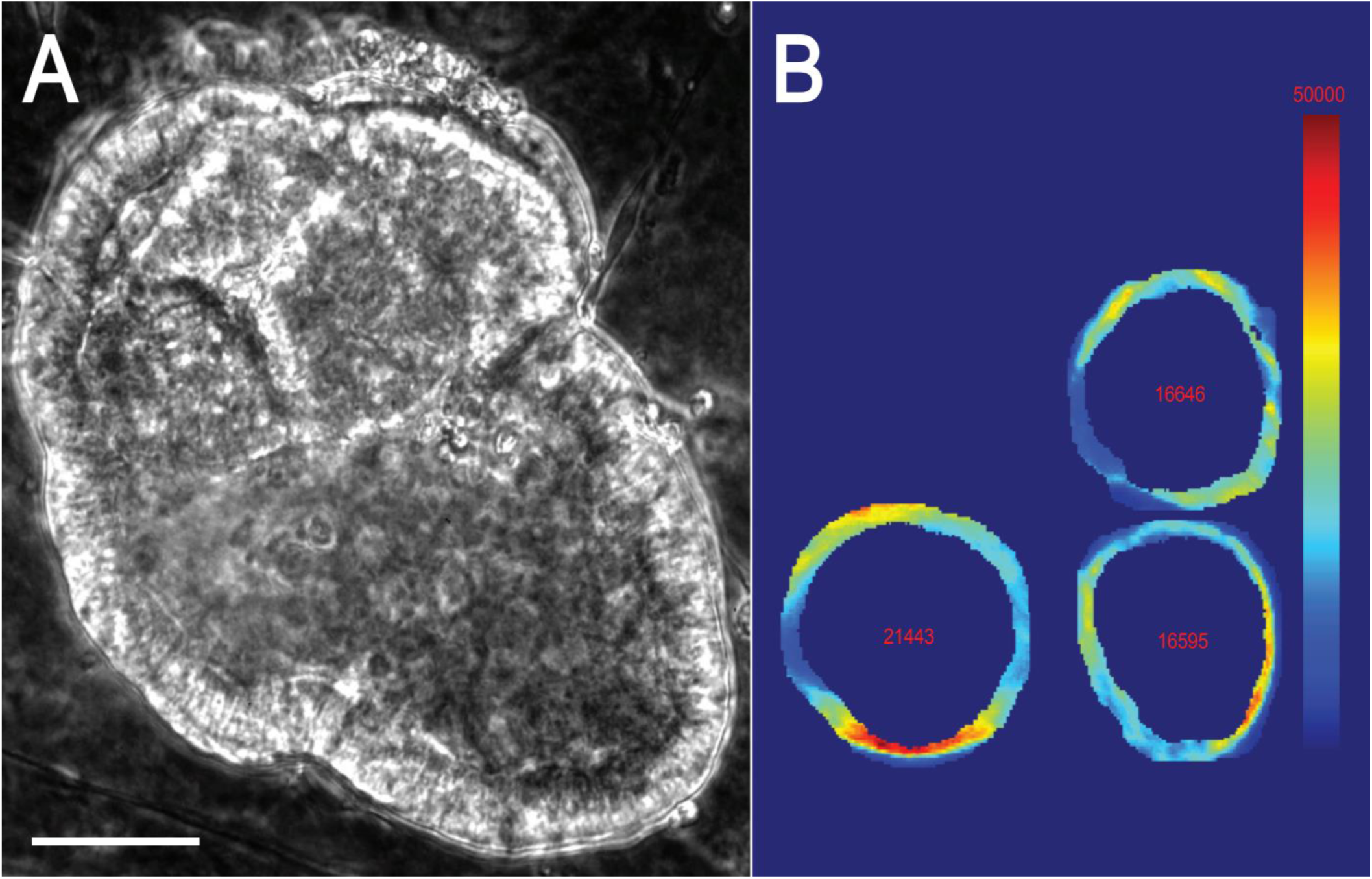
Metastatic prostate cancer organoids obtained from micro-metastasis at a retroperitoneal lymph node. (A) Organoids derived from radical prostatectomy tissue. Day 105. Organoid whole genome sequencing showed homozygous mutations in *PTEN* and *BRCA2*. Phase contrast microscopy, magnification 20x. Scale bar equals 50 µm. (B) Pixel intensity of MT bundles in A549 lung cancer cells treated with 100 nM paclitaxel.

**Table S1.** Statistical analysis of MT morphology and localization in diffuse gastric cancer cells. 50 cells from each of the 7 gastric cancer cell lines were treated with 100 nM docetaxel. Changes in the morphology and localization of the MT cytoskeleton was evaluated based on 21 numerical metrics in immunofluorescence images. Statistical significance (highlighted in red) was determined based on a Student’s *t*-test.

| Cell Line | SCH |  |
| --- | --- | --- |
| FeatureName | p < 0.01 | p |
| object:number | 0 | 0.3666 |
| object:EulerNumber | 0 | 0.0468 |
| object_size:average | 0 | 0.9385 |
| object_size:variance | 0 | 0.1852 |
| object_size:ratio | 0 | 0.053 |
| object_distance:average | 0 | 0.1785 |
| object_distance:variance | 0 | 0.0292 |
| object_distance:ratio | 0 | 0.7446 |
| edges:area_fraction | 0 | 0.0177 |
| edges:homogeneity | 0 | 0.9207 |
| edges:direction_maxmin_ratio | 0 | 0.1135 |
| edges:direction_maxnextmax_ratio | 1 | 0.0073 |
| edges:direction_difference | 0 | 0.2082 |
| obj_skel_len | 0 | 0.5825 |
| obj_skel_hull_area_ratio | 0 | 0.1697 |
| obj_skel_obj_area_ratio | 0 | 0.1122 |
| obj_skel_obj_fluor_ratio | 0 | 0.0993 |
| obj_skel_branch_per_len | 0 | 0.2128 |
| convex_hull:fraction_of_overlap | 0 | 0.0164 |
| convex_hull:shape_factor | 1 | 0.0015 |
| convex_hull:eccentricity | 1 | 0.0004 |
| Cell Line | HS746T |  |
| FeatureName | p < 0.01 | p |
| object:number | 0 | 0.4485 |
| object:EulerNumber | 0 | 0.0646 |
| object_size:average | 0 | 0.6876 |
| object_size:variance | 0 | 0.5237 |
| object_size:ratio | 0 | 0.0279 |
| object_distance:average | 0 | 0.3173 |
| object_distance:variance | 0 | 0.2418 |
| object_distance:ratio | 0 | 0.8296 |
| edges:area_fraction | 1 | 0.0011 |
| edges:homogeneity | 0 | 0.7999 |
| edges:direction_maxmin_ratio | 1 | 0.0004 |
| edges:direction_maxnextmax_ratio | 0 | 0.4087 |
| edges:direction_difference | 0 | 0.1352 |
| obj_skel_len | 0 | 0.546 |
| obj_skel_hull_area_ratio | 0 | 0.3784 |
| obj_skel_obj_area_ratio | 0 | 0.9464 |
| obj_skel_obj_fluor_ratio | 0 | 0.9765 |
| obj_skel_branch_per_len | 0 | 0.457 |
| convex_hull:fraction_of_overlap | 0 | 0.3399 |
| convex_hull:shape_factor | 0 | 0.3004 |
| convex_hull:eccentricity | 0 | 0.8351 |
| **Cell Line** | **MKN7** |  |
| FeatureName | p < 0.01 | p |
| object:number | 0 | 0.5177 |
| object:EulerNumber | 0 | 0.1326 |
| object_size:average | 0 | 0.089 |
| object_size:variance | 0 | 0.0159 |
| object_size:ratio | 0 | 0.0283 |
| object_distance:average | 1 | 0 |
| object_distance:variance | 0 | 0.3068 |
| object_distance:ratio | 0 | 0.1857 |
| edges:area_fraction | 0 | 0.2718 |
| edges:homogeneity | 0 | 0.0659 |
| edges:direction_maxmin_ratio | 0 | 0.2596 |
| edges:direction_maxnextmax_ratio | 0 | 0.0596 |
| edges:direction_difference | 0 | 0.1094 |
| obj_skel_len | 0 | 0.0373 |
| obj_skel_hull_area_ratio | 0 | 0.7156 |
| obj_skel_obj_area_ratio | 0 | 0.117 |
| obj_skel_obj_fluor_ratio | 0 | 0.1162 |
| obj_skel_branch_per_len | 0 | 0.0145 |
| convex_hull:fraction_of_overlap | 0 | 0.2667 |
| convex_hull:shape_factor | 0 | 0.0687 |
| convex_hull:eccentricity | 0 | 0.6528 |
| **Cell Line** | **TMK1** |  |
| FeatureName | p < 0.01 | p |
| object:number | 1 | 0.0004 |
| object:EulerNumber | 1 | 0 |
| object_size:average | 1 | 0 |
| object_size:variance | 1 | 0 |
| object_size:ratio | 0 | 0.0181 |
| object_distance:average | 0 | 0.0489 |
| object_distance:variance | 1 | 0.0001 |
| object_distance:ratio | 0 | 0.0528 |
| edges:area_fraction | 0 | 0.0135 |
| edges:homogeneity | 1 | 0 |
| edges:direction_maxmin_ratio | 1 | 0.009 |
| edges:direction_maxnextmax_ratio | 0 | 0.1082 |
| edges:direction_difference | 0 | 0.9562 |
| obj_skel_len | 1 | 0 |
| obj_skel_hull_area_ratio | 0 | 0.2486 |
| obj_skel_obj_area_ratio | 0 | 0.4571 |
| obj_skel_obj_fluor_ratio | 0 | 0.4648 |
| obj_skel_branch_per_len | 0 | 0.6773 |
| convex_hull:fraction_of_overlap | 1 | 0 |
| convex_hull:shape_factor | 1 | 0.0081 |
| convex_hull:eccentricity | 1 | 0.0004 |
| **Cell Line** | **SNU1** |  |
| FeatureName | p < 0.01 | p |
| object:number | 0 | 0.2043 |
| object:EulerNumber | 0 | 0.0279 |
| object_size:average | 0 | 0.039 |
| object_size:variance | 0 | 0.1368 |
| object_size:ratio | 0 | 0.3181 |
| object_distance:average | 0 | 0.2248 |
| object_distance:variance | 0 | 0.9796 |
| object_distance:ratio | 0 | 0.6974 |
| edges:area_fraction | 1 | 0 |
| edges:homogeneity | 0 | 0.0435 |
| edges:direction_maxmin_ratio | 1 | 0.0021 |
| edges:direction_maxnextmax_ratio | 1 | 0.0003 |
| edges:direction_difference | 1 | 0.008 |
| obj_skel_len | 0 | 0.1965 |
| obj_skel_hull_area_ratio | 0 | 0.2808 |
| obj_skel_obj_area_ratio | 0 | 0.1603 |
| obj_skel_obj_fluor_ratio | 0 | 0.1551 |
| obj_skel_branch_per_len | 0 | 0.8024 |
| convex_hull:fraction_of_overlap | 0 | 0.2285 |
| convex_hull:shape_factor | 0 | 0.5757 |
| convex_hull:eccentricity | 0 | 0.6879 |
| **Cell Line** | **AZ521** |  |
| FeatureName | p < 0.01 | p |
| object:number | 0 | 0.1103 |
| object:EulerNumber | 0 | 0.0785 |
| object_size:average | 0 | 0.0254 |
| object_size:variance | 0 | 0.1188 |
| object_size:ratio | 0 | 0.0263 |
| object_distance:average | 0 | 0.0692 |
| object_distance:variance | 0 | 0.5256 |
| object_distance:ratio | 0 | 0.42 |
| edges:area_fraction | 1 | 0 |
| edges:homogeneity | 0 | 0.1146 |
| edges:direction_maxmin_ratio | 0 | 0.0136 |
| edges:direction_maxnextmax_ratio | 0 | 0.0723 |
| edges:direction_difference | 0 | 0.1317 |
| obj_skel_len | 1 | 0.0038 |
| obj_skel_hull_area_ratio | 1 | 0.0003 |
| obj_skel_obj_area_ratio | 0 | 0.6782 |
| obj_skel_obj_fluor_ratio | 0 | 0.8323 |
| obj_skel_branch_per_len | 0 | 0.9507 |
| convex_hull:fraction_of_overlap | 1 | 0.0001 |
| convex_hull:shape_factor | 1 | 0.0063 |
| convex_hull:eccentricity | 0 | 0.1111 |
| **Cell Line** | **MKN45** |  |
| FeatureName | p < 0.01 | p |
| object:number | 0 | 0.386 |
| object:EulerNumber | 0 | 0.0151 |
| object_size:average | 1 | 0.0004 |
| object_size:variance | 0 | 0.0234 |
| object_size:ratio | 0 | 0.554 |
| object_distance:average | 0 | 0.6923 |
| object_distance:variance | 0 | 0.452 |
| object_distance:ratio | 0 | 0.4829 |
| edges:area_fraction | 0 | 0.0285 |
| edges:homogeneity | 1 | 0.0036 |
| edges:direction_maxmin_ratio | 0 | 0.6929 |
| edges:direction_maxnextmax_ratio | 0 | 0.2949 |
| edges:direction_difference | 0 | 0.5692 |
| obj_skel_len | 1 | 0.0023 |
| obj_skel_hull_area_ratio | 0 | 0.73 |
| obj_skel_obj_area_ratio | 0 | 0.7947 |
| obj_skel_obj_fluor_ratio | 0 | 0.7844 |
| obj_skel_branch_per_len | 0 | 0.2782 |
| convex_hull:fraction_of_overlap | 0 | 0.1549 |
| convex_hull:shape_factor | 0 | 0.7487 |
| convex_hull:eccentricity | 0 | 0.3172 |

Video 1 – Prostate cancer organoid 72 hours imaging. Organoids were seeded after prostate tumor cells were derived by tissue dissociation of radical prostatectomy samples. https://vimeo.com/1063891309/bb904d8da5

